# Semi-Random Mixing Epidemic Model: Integrating Explicit Household and Non-household Interactions

**DOI:** 10.1101/2025.11.13.25340112

**Authors:** Michael Lazarus Smah, Anna C. Seale, Kat S. Rock

**Affiliations:** Institute for Global Pandemic Planning, Warwick Medical School, Coventry, CV4 7AL, United Kingdom; System Biology and Infectious Disease Epidemiology Research, Warwick Mathematics Institute, Coventry, CV4 7AL, United Kingdom; London School of Hygiene & Tropical Medicine

## Abstract

Understanding how infectious diseases spread through populations requires models that capture real human interactions more realistically. Many classical epidemic models assume that everyone mixes randomly, overlooking the structured and clustered nature of daily contacts within households, workplaces, and schools. These simplifications can limit their predictive capability and the design of effective control strategies. To improve on this limitation, we develop an epidemic modelling framework that integrates explicitly both household and non-household interactions. Building on the semi-random mixing (SeRaMix) concept in the literature, the model captures how people move between different major contact settings (household and work/school) each day, interacting within and across clusters of contacts. We introduce a novel formulation linking contact duration and proximity to infection risk, enabling a more realistic representation of disease-specific transmission factors in an equation-based model. Analytical derivation of the basic reproduction number (*R*_0_) demonstrates how epidemic potential distinctly depends on social behaviour, mobility, and biological parameters. Validation against an individual-based model confirms that this equation-based framework reproduces epidemic dynamics within this structured network of interaction. Sensitivity analyses identify the number of contacts at home and work/school, mobility, and inter-household connections as key non-pathogen-dependent drivers of epidemic growth. This framework offers a more realistic approach to analysing the impacts of non-pharmaceutical interventions—such as reduced mobility, hybrid working, and household bubbles—on outbreak trajectories.

## 1 Background

Human susceptibility, mobility, and number of contacts, as well as the infectiousness of the pathogen, influence infectious disease outbreaks and their propensity to become epidemics or pandemics. The number of usual contacts and their duration and proximity between them may depend on whether interactions are within or outside the residence (typically a household but could be a larger institution) and are likely different from those outside (e.g., work and school). However, these differing factors and mobility are rarely completely accounted for in modelling infectious disease spread, and this may impact modelling analyses of non-pharmaceutical interventions (NPIs) directed to reduce human mobility and interactions.

Capturing the critical aspects of human mobility and interactions explicitly in epidemic models may improve modelling insights into epidemic growth and consider how changing these human interactions, through NPIs, might affect the course of an outbreak. In most simple classical epidemic models, the effects of mobility, number of contacts, and local population size are often described with a single parameter—the effective contact rate (contact rate modification to include susceptibility, behavioural factors, and pathogen characteristics). In addition, the distinction between interactions at home and those outside is sometimes ignored. The simplified assumptions reduce complexities in modelling analyses but may lead to misleading results in assessing potential NPIs [1, 2].

Capturing household interactions in epidemic models is complex, although considerable efforts have been made, particularly in recent years [3–5]. Equation-based models that incorporate household structure are usually characterised by large equations [6], and do not explicitly account for the mobility that links the household and the non-household interactions, thereby increasing the complexity of their use.

We proposed the concept of *semi-random mixing* (SeRaMix) in *a potentially recurrent (multi-clique) contact network* [7] to contribute to the understanding of how incorporating clustered interactions in the force of infection may affect the prediction of epidemic trajectories. We proposed the SeRaMix framework to account for cluster-based interactions explicitly in the classical epidemic models that relax the simplifying assumption of random mixing. An extension of this approach offers a way to capture interactions in transmission that is suitable for both household and non-household contacts, as both household and non-household interactions are characterised by multiple cluster formation rather than interaction with the general population, as in a single cluster; in this study, we considered that such random contact mixing occurs locally within clusters of interaction. In [7], we proposed that everyone has specific potential contacts (a subset of the entire population) from which they randomly choose their actual contacts at each time step. Potential contacts are drawn from an individual’s cluster (household, workplace, or school). This relationship results in increased infection spread in populations with larger cluster sizes compared to those with smaller cluster sizes.

The SeRaMix model introduced in [7] supports the ideas in the literature [8], which suggest that interactions are largely local rather than globally random. This is because most individuals are likely to only have random contacts locally. After all, they have the same set of people who define local interaction (e.g., household members, schoolmates, colleagues at work, and other external neighbours) from whom daily contacts may occur. We proposed a modification to the force of infection in classical epidemiological models to include dependence on the average potential contacts per time per person, which includes contacts with members of one’s cluster and inter-cluster contacts.

The idea of incorporating two levels of interaction in a single model has been explored in the literature [9, 10] and accounts for the probability of infection per unit time as a sum of typical daytime and nighttime interactions. These models use the metapopulation approach, which models the spread of infection across different locations. They take into account the probability of infection when individuals move to work (day), which includes interaction between individuals from different locations who have moved to work/school, and the probability of infection when they return home (night), which includes interaction between members of the same location. However, they do not include cluster/household structures that distinguish between household and non-household interactions.

In this study, we built on the work of [7], now incorporating concepts of “Movement-Interaction-Return” [10] in a discrete-time SeRaMix model that accounts for different mixing within and outside the households. However, the specific locations where people interact during non-household interactions will not be explicitly incorporated. This is to allow a focus on the adaptation of the cluster-based interaction in [7] for multi-level interaction epidemic modelling. Because of the multi-level interactions, we propose a framework to incorporate the differences in the contact duration and proximity between contacts within and outside the household.

## 2 Methods

In this study, household and non-household interactions differ in terms of cluster sizes, external connectivity, contact duration, and proximity. Here, we consider mobility originating from the residence/household after household/home-time interaction. Thus, the order is *Home interaction - Mobility - Work interaction - Return*. This captures how household clusters are formed during home-time interaction, with household members dispersing during work time, essentially shuffling household members, with each going to different work/school clusters and interacting with different people before returning home. The aim here is to accommodate different cluster sizes, contact duration, and proximity for household and non-household interactions in the spread of infectious diseases, and how these impact our understanding of epidemic progress and control.

This study will begin by proposing a method for accounting for contact duration and proximity between contacts before formulating the multi-level interaction model (household and non-household interactions).

### 2.1 Modelling contact duration and proximity

Research on the risk of transmission of SARS-CoV-2 (causing COVID-19) revealed that the duration and proximity of exposure to an infectious person were key determinants of the transmission risk, with the risk of transmission increasing with an increase in the duration of close exposure between contacts [11]. Efforts have been made in the literature to quantify the contact duration and distance between contacts required for transmission of infectious pathogens [12–16]. These quantities are likely to have different values for different diseases, as illustrated in Table 1, which gives four examples of diseases and their definitions of what constitutes a close contact that may lead to transmission. It is important to consider in this modelling framework how we may incorporate these metrics.

**Table 1.**
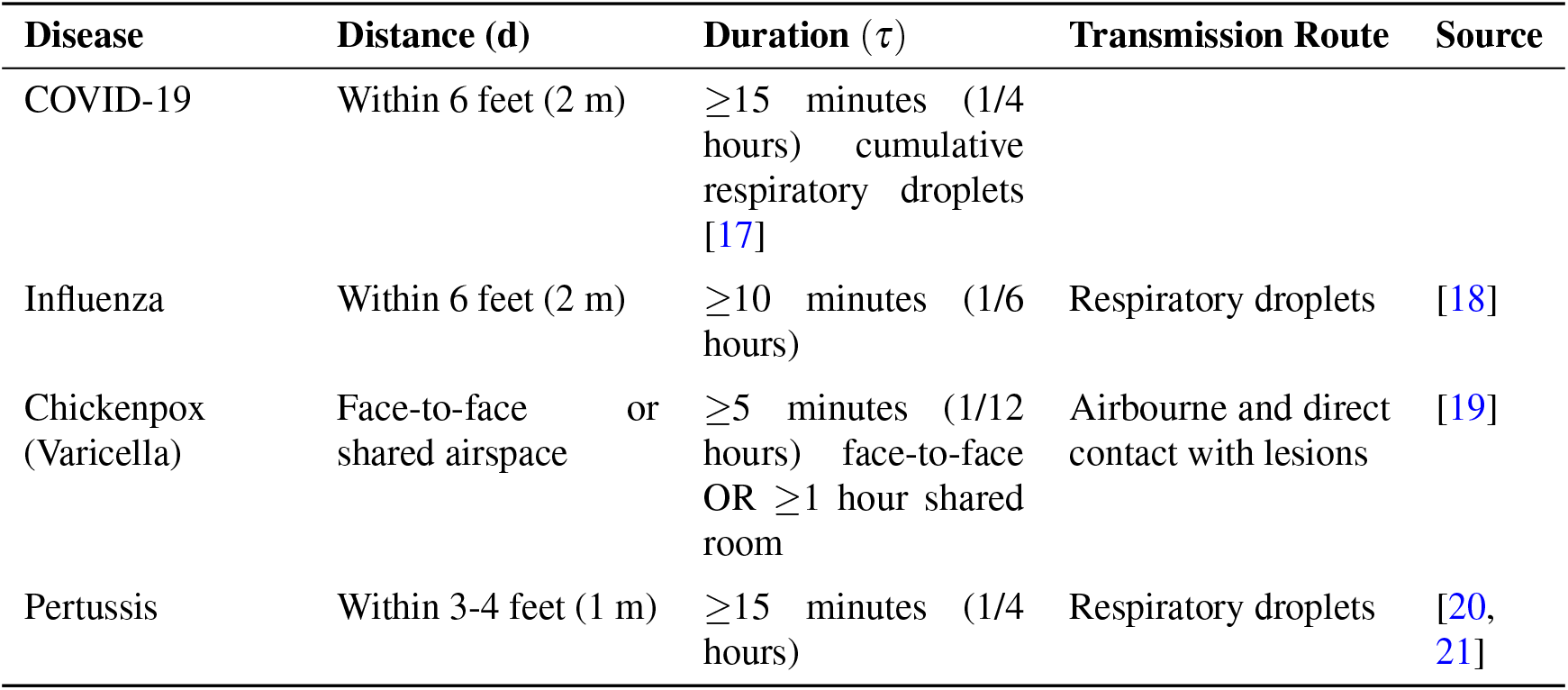
Examples of “close contact” definitions for a range of infectious pathogens compiled from the literature. The values given are the threshold distance and duration for infection to be considered likely.

| Disease | Distance (d) | Duration ($\tau$) | Transmission Route | Source |
| --- | --- | --- | --- | --- |
| COVID-19 | Within 6 feet (2 m) | $\geq 15$ minutes (1/4 hours) cumulative respiratory droplets [17] | | |
| Influenza | Within 6 feet (2 m) | $\geq 10$ minutes (1/6 hours) | Respiratory droplets | [18] |
| Chickenpox (Varicella) | Face-to-face or shared airspace | $\geq 5$ minutes (1/12 hours) face-to-face OR $\geq 1$ hour shared room | Airbourne and direct contact with lesions | [19] |
| Pertussis | Within 3-4 feet (1 m) | $\geq 15$ minutes (1/4 hours) | Respiratory droplets | [20, 21] |

Here, we propose the use of a modified sigmoid function to account for the impacts of contact duration and proximity between contacts in models of the effective contact size. Suppose that the threshold contact duration required for an infection to be transmitted is given by *τ*, and the threshold distance between contacts to transmit infection is *d*. If the average contact duration in the population is *T*, and the average distance between contacts is *D*. Then we assume that the risk of transmission increases with the increase in *Effective contact size* defined by:

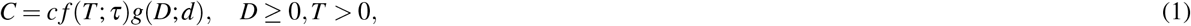

where 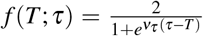, is the Duration Risk index represents the effect of contact duration on the transmission and 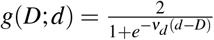, is the Proximity Risk index representing the effect of contact proximity on the transmission, *c* ≥ 1 is the average number of contacts per unit of time, *ν*_*τ*_ is the steepness parameter that represents the speed of effective contact duration, and *ν*_*d*_ is the steepness parameter representing the speed of effective contact distance. The steepness parameters control how sharply the functions ( *f* (*T* ; *τ*) and *g*(*D*; *d*)) transition from low to high around the midpoint. As T increases, the chance of transmission increases, and as D increases, the chance of transmission decreases.

The functions *f* (*T* ; *τ*) and *g*(*D*; *d*) have been constructed by modifying the sigmoid function [22, 23] such that a transition exists around the threshold required for an infection to be transmitted through person-to-person contact. The behaviour of these functions are analysed and visualised in the Supplementary Information.

### 2.2 Modelling household and non-household interactions

To formulate the multi-stage interactions and infection, we use the steps in **Algorithm 1** (Supplementary Information). First, we consider that at each time step, a proportion *α* ≥ 0 of people will leave their homes for work/school, while a proportion (1 − *α*) will remain home, where non-work-related interaction may occur. Let *S*(*t*), *I*(*t*), *R*(*t*) be the numbers of the susceptible, infected, and recovered (removed) individuals at time *t*, respectively. We assume that all individuals return home after work-time interactions and are at risk of being infected at home during the home-time interaction through the household probability of infection. Furthermore, we assume that during work-time interactions, any susceptible person who does not move to work remains and interacts with other people at home and can only be infected by infected persons who do not go to work. Susceptible individuals who go to work have the risk of being infected during work-time interaction outside their households. We assume that infections that occur within household areas after the return stage and before the next movement stage are counted as infections of the next time step occurring during home-time interaction. Thus, the daily multi-level infection begins with the home-time infection before the movement stage and ends when people return home (see the Supplementary Information for detailed derivation).

The derived total FOI on day *t* is given by:

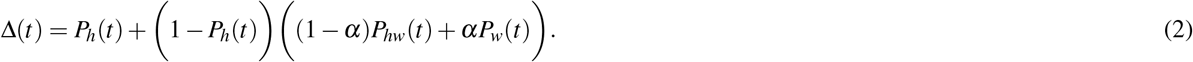

Using a similar approach in [7], we can compute the *P*_*h*_(*t*), *P*_*hw*_(*t*), and *P*_*w*_(*t*) as:

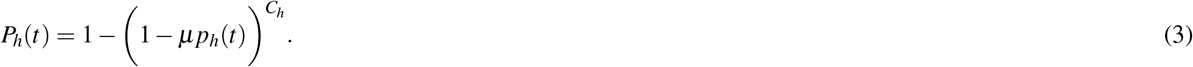

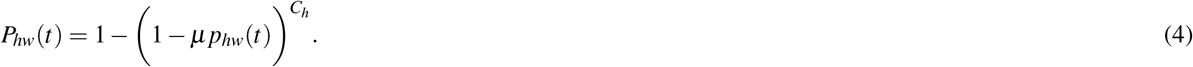

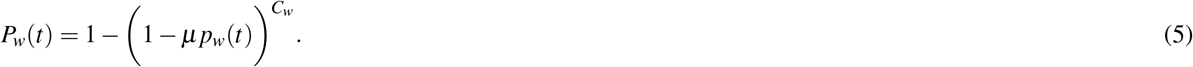

The terms *p*_*h*_(*t*), *p*_*hw*_(*t*), and *p*_*w*_(*t*) are given by:

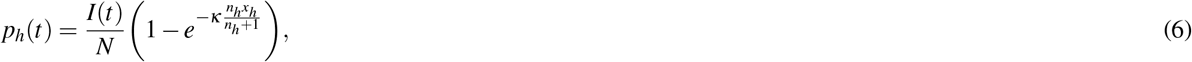

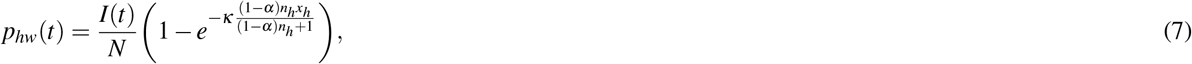

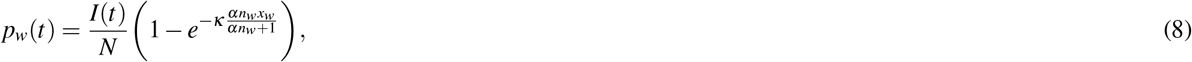

where *µ* is the pathogen-dependent probability that infection will be transmitted after contact with an infected person, and *C*_*h*_ is the average household contact size, and *C*_*w*_ is the average work contact size.

The household *effective contact size* is given by:

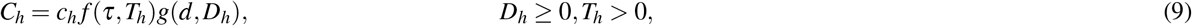

where *c*_*h*_ is the average number of household contacts, *T*_*h*_ is the average household contact duration, and *D*_*h*_ is the average household contact proximity, and the work *effective contact size* is given by:

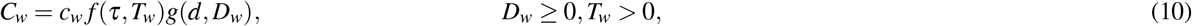

where *c*_*w*_ is the average work contact, *T*_*w*_ is the average contact duration, and *D*_*w*_ is the average contact distance outside the residence/household.

Given that the infected individual recovers with probability *γ*, the discrete-time Markovian equations for the SIR-type infection are given as:

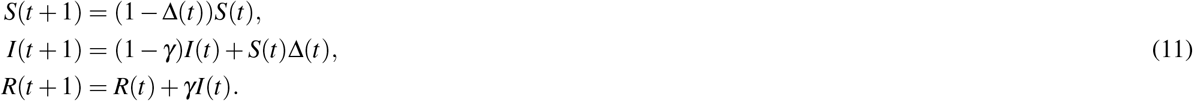

Where *S*(*t*), *I*(*t*), *R*(*t*) are the numbers of the susceptible, infected, and recovered (removed) individuals at time *t* respectively, and *S*(*t*) + *I*(*t*) + *R*(*t*) = *N*, is the total population.

### 2.3 Model validation and analysis

An individual-based modelling (IBM) approach can provide further insight into epidemic spread, complementing equation-based models (EBMs) by capturing heterogeneous mixing patterns, small-scale clustering effects, and stochastic variability in early outbreaks. The framework enables systematic investigation of how household/workplace sizes (*n*_*h*_, *n*_*w*_), a measure of mobility (*α*), contact rates (*c*_*h*_, *c*_*w*_) and external connectivity (*x*_*h*_, *x*_*w*_) collectively influence epidemic trajectories and transmission patterns.

Similar to the approach in the literature [7], our analogous IBM (**Algorithms** 2–6) (Supplementary Information) is used here to test and compare the results of the EBM developed in this study.

The simulation implements an IBM to capture disease transmission dynamics through household and workplace interactions, incorporating daily mobility patterns. The process begins with the initialisation of a population of *N* = 1000 individuals, organised into household clusters of size *n*_*h*_ and workplace clusters of size *n*_*w*_. To construct the household network, *N* individuals are randomly assigned to clusters of size *n*_*h*_. For each individual, the cluster neighbours are assigned as primary neighbours. External neighbours are randomly assigned from individuals from other households. Each individual is assigned the indices of their primary and external neighbours from which daily contacts could be selected. The same process is done for the cluster assignment, such that cluster neighbours and external neighbours at work/school are assigned regardless of whether people were from the same households (details of the network construction are in **Algorithms 2–3** (Supplementary Information).

The epidemic is seeded by randomly assigning a single infected individual, with all others initially susceptible. Health states are numerically encoded as susceptible (*S* = 0), infected (*I* = 1), or removed/recovered (*R* = 2). Daily progression follows a structured sequence of transmission phases. First, household transmission before mobility occurs as susceptible individuals face infection risk through random selection of *c*_*h*_ household contacts, with infection probability *µ* per infectious contact. This phase captures home-time household transmission through both core household members and external household connections.

Subsequently, each agent independently decides to stay home (probability 1 − *α*) or go to work (probability *α*), creating dynamically changing subpopulations. The household transmission phase then continues for the stay-at-home group, where non-mobile susceptible individuals experience infection risk only from other stay-at-home household members and external connections, maintaining the same contact rate *c*_*h*_. This model configuration ensures an increase in the risk of home transmission when family members remain together throughout the day without going out to work/school.

Mobile individuals instead face workplace transmission risks through random selection of *c*_*w*_ work contacts, with the same per-contact infection probability *µ* for each effective contact (close enough and for sufficient contact duration). This occupational exposure occurs through both core workplace members and external work connections. Infected individuals recover with probability *γ* each day, transitioning to the permanently immune removed state.

Simulation runs continue until either reaching the maximum simulation time or disease extinction. Only realisations meeting a pre-defined outbreak threshold are retained, filtering out stochastic fade-outs to focus on established epidemics (for better comparison with the deterministic EBM). The model tracks time-series counts of susceptible, infected, and removed individuals, while distinguishing infection sources as home-before-mobility, home-after-mobility, and workplace transmissions. Results aggregate across multiple network configurations for varying *x*_*h*_, *x*_*w*_ and realisations.

Except where indicated, the baseline parameter values in Table 2 have been used for simulation.

The basic reproduction number is given as:

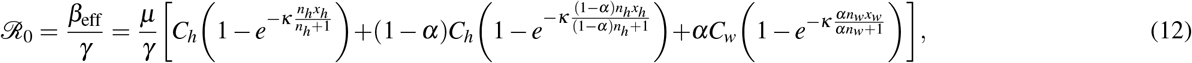

where the effective transmission rate is:

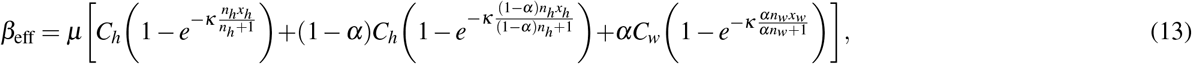

**Table 2.**
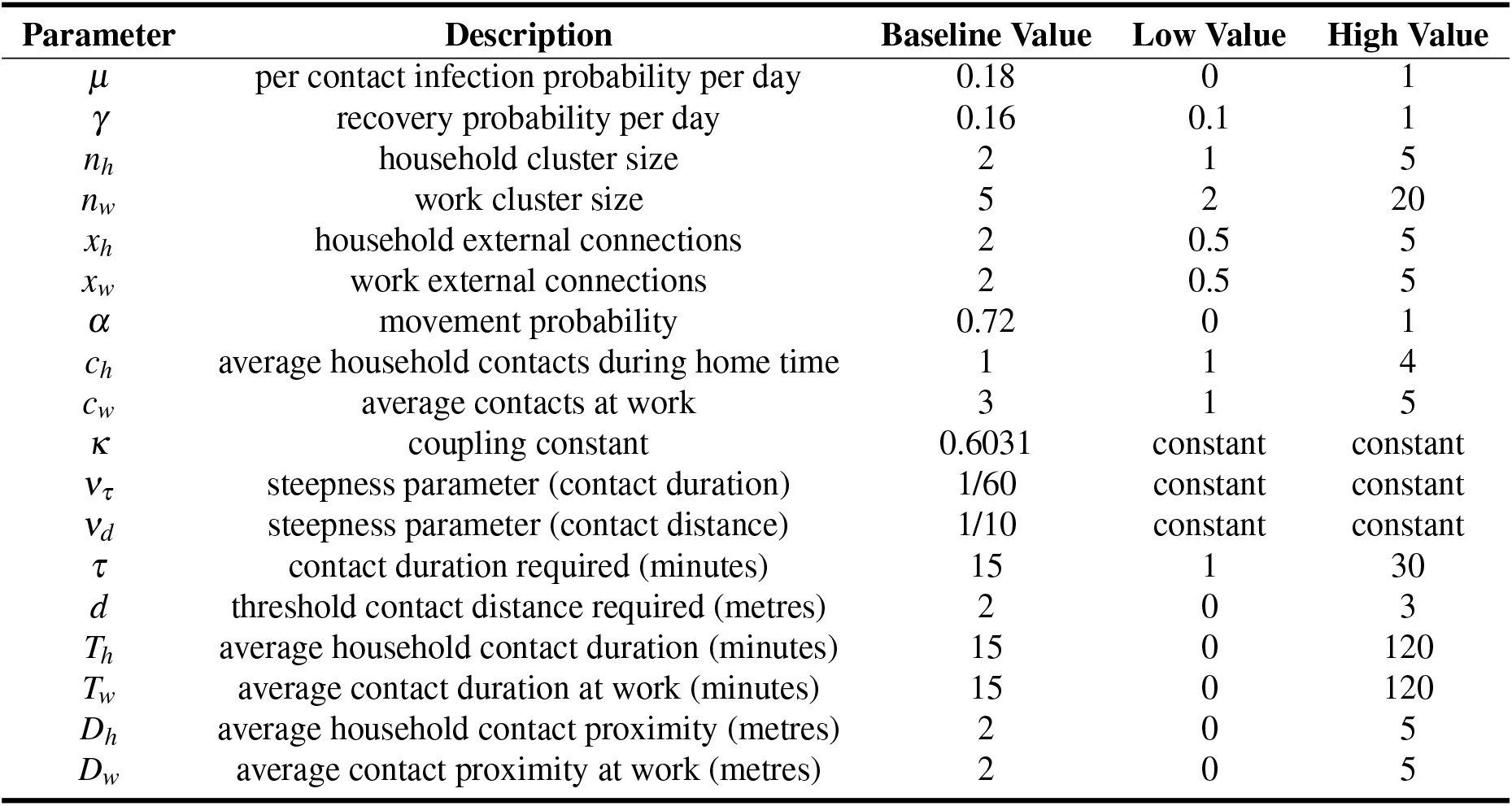
Parameters description, baseline values, and their ranges. These parameters are selected to be illustrative of a COVID-like infection; however, they are intended as exemplar parameterisation rather than reflecting a specific pathogen and population.

Substituting for *C*_*h*_, and *C*_*w*_:

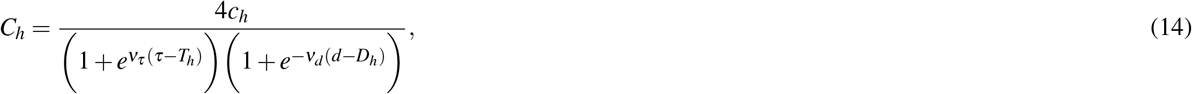

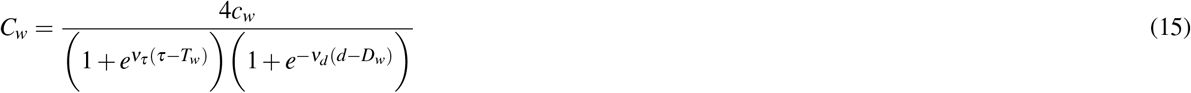

gives

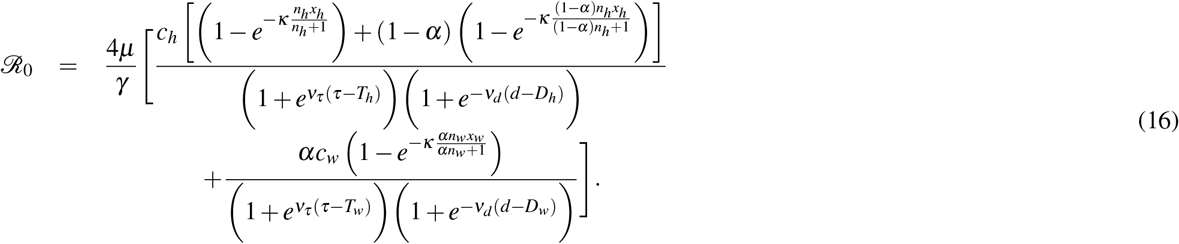

### 2.4 Sensitivity analysis of ℛ_0_

The sensitivity analysis of ℛ_0_ is presented in this section to deduce the impact of each parameter on ℛ_0_. The local sensitivity analysis, which is a one-at-a-time (OAT) technique to find the influence of each parameter while keeping other parameters fixed [24], uses computation of partial derivatives of ℛ_0_ with respect to each parameter 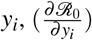, to compute the sensitivity indices using the relationship

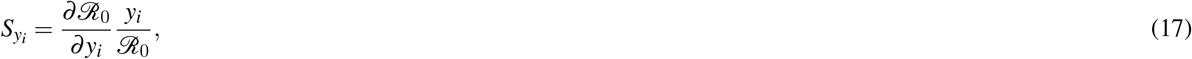

which is the normalised sensitivity index, which expresses the relative change in ℛ_0_ due to a relative change in *y*_*i*_.

A positive 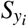 signifies that the function value (in this model, ℛ_0_) will increase as the parameter value *y*_*i*_ increases, while a negative 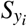 means that the function value will decrease as the parameter *y*_*i*_ increases [25]. The analytical expressions of the partial derivatives computed above do not communicate to us the order of importance in the sensitivity indices of the parameters given their default values and ranges; as such, numerical results are presented in the next section.

### 2.5 Numerical simulation

We performed numerical simulations to present the numerical results of the sensitivity analysis (local and global) and the effect of parameter reduction on the dynamics of infectious diseases. Except where indicated, the values of the baseline parameters used for numerical simulation are in Table 2 corresponding to ℛ_0_ = 2.22. Since the biological infectiousness and recovery are probabilities, the lower and upper bounds are set as *µ* ∈ [0, 1] and, to avoid errors with division by zero, the recovery probability range is *γ*∈ [0.1, 1]. Based on the report that 80% of the countries in the world have household sizes ranging from 2.3–5 [26], the range of household size is set to *n*_*h*_∈ [1, 5]. Based on this, the household contact range is set as *c*_*h*_ ∈ [1, 4]. The proportion of people moving from home/households to work/school is given by *α* ∈ [0, 1]. Details of baseline other parameter values and their possible range (as assumed) are in Table 2. For the EBM simulation, we set initial values of the variables *N* = 1, 000, *I*(0) = 1, *R*(0) = 0, *S*(0) = *N* − *I*(0) − *R*(0).

A global sensitivity analysis was performed to ascertain the proportion of the global effect that can be attributed to each parameter on the ℛ_0_. Since it is unfeasible to be able to influence the biological characteristics of pathogens, we wanted to explore how parameters that can be changed through efforts to control the outbreak modify ℛ_0_. Therefore, we set the biological parameters *µ, γ, τ*, and *d*, and the coupling parameter *κ* to be constant while performing the global sensitivity analysis on only the non-biological parameters. Although *γ* is simply considered a biological parameter here, it is worth stating that, in the SIR model (as presented in this study), *γ* is the probability (rate for a continuous-time model) of recovery or removal, which tells us how rapidly infected people leave the state of being infected and capable of transmitting infection. Antivirals are examples of pharmaceutical interventions that can increase the removal of infected people by making infections last less time. Similarly, NPIs, including testing, symptom-based detection, and isolation, also increase the effective removal rate since people who have been removed cannot spread the disease any further. Using only the biological parameters also helps reduce the number of parameters and allows clear visualisation of the decision variables. Latin hypercube sampling (LHS) is used to draw 10,000 samples for each of the nonbiological parameters of the model (in the range of values in Table 2), resulting in a 10,000 by 11 matrix, where each row defines a set of unique parameters. The LHS was first introduced by McKay et al. [27] and is a statistical technique for producing a nearly random sample of parameter values from a multidimensional parameter space. It is a stratified sampling method that splits the range of each parameter into 𝒩 equally likely intervals and makes sure that only one sample is taken from each interval for each parameter. A set of 𝒩 points in the parameter space is then created by randomly combining the samples across parameters. Compared to random sampling, this approach ensures that the entire range of each parameter is explored, which reduces the likelihood of clustering and provides good coverage [28, 29]. The rows were used to calculate the response functions, ℛ_0_. In this analysis, rather than varying the parameters one at a time, the input parameters are varied simultaneously using the unique set of parameters generated, thereby computing the relative contribution of each parameter and its interaction with other parameters in the variance of the model output [30].

The Sobol Global Sensitivity Analysis method [30, 31] was used. In this approach, the steps in **Algorithm 7** (Supplementary Information) were followed to implement the sensitivity analysis in MATLAB:

The total variance *V* is calculated as:

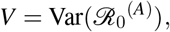

where ℛ_0_^(*A*)^ is the model output evaluated using the parameter samples for each *c* ∈ *C*. This is a measure of how “spread out” the model outputs are, which quantifies how much uncertainty exists in the model output due to the variation in all input parameters. The first-order variance contribution *V*_*i*_ captures the variance in the output that is solely due to parameter *i*, without considering interactions with other parameters. It is calculated as:

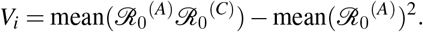

Where ℛ_0_^(*A*)^ represents the model output when a set of parameters *A* is used. ℛ_0_^(*C*)^ represents the model output when only one parameter *i* is replaced by its counterpart in set *B*, while all other parameters remain the same. The term mean(ℛ_0_^(*A*)^ℛ_0_^(*C*)^) captures how much the outputs of *A* and *C* are correlated due to parameter *i*. Subtracting mean(ℛ_0_^(*A*)^)^2^ removes the baseline variance that arises purely from the mean output value, leaving only the variance contribution from parameter *i*. The total-order variance contribution 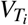 captures all the variance in the output that involves parameter *i*, including its interactions with other parameters. It is calculated as:

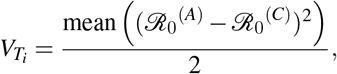

where (ℛ_0_^(*A*)^ − ℛ_0_^(*C*)^)^2^ measures the squared difference in output due to changing parameter *i*. The mean of these squared differences represents how much variance is introduced by *i*, including any interactions *i* has with other parameters. Dividing by 2 adjusts for the fact that the squared difference includes contributions in both directions (from *A* to *C* and vice versa).

## 3 Results

The analytical expression of the basic reproduction number derived using this framework was given in the methods section (equation (16)). It showed that, in addition to dependence on the recovery and disease infectious probabilities, the way in which (ℛ_0_) is dependent on the proportion of people moving to work, *α*, the average household and work-time number of contacts, *C*_*h*_ and *C*_*w*_, the average household/household size *n*_*h*_, the average work-time cluster size *n*_*w*_, and the average external connections at home, *x*_*h*_, and at work, *x*_*w*_. This allows us to analyse the impacts of the parameters on the ℛ_0_ and analyse NPIs targeting different aspects of movement and interactions (household and non-household) captured in this model.

The different epidemic trajectories in Figure 1 reveal how different contact phases (home or work/school) contribute to epidemic spread under varying external connection scenarios (indicated by *x*_*h*_ and *x*_*w*_ values) as captured by the IBM simulation, where the number of contacts remains the same across a varying pool of potentially recurrent contacts. Epidemic growth is dominated by work/school transmission, which is seen as the larger amplitudes of new infections of the work/school transmission phase across all pairs of connections, *x*_*h*_ and *x*_*w*_. New infections increase as external connectivity increases, as the increase in external connectivity increases the pool of potentially recurrent contacts for each individual, although the same average contacts are randomly selected at each time step.

**Figure 1.**
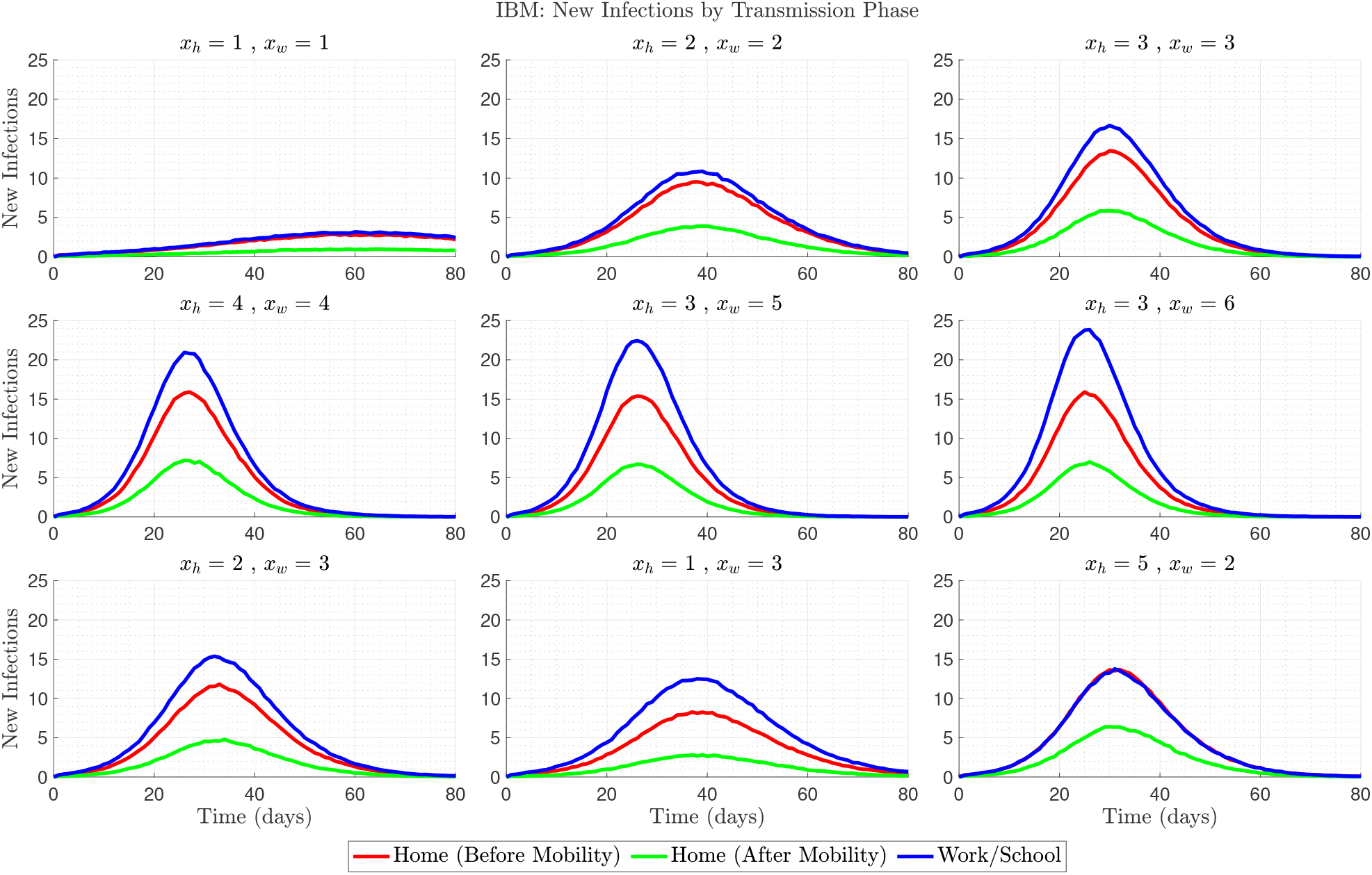
IBM simulation results showing daily new infections (average of 1,000 realisations) stratified by transmission phase: home before mobility (red), home after mobility (green), and workplace (blue).

Figures 2 and 3 show that the EBM dynamics are well within a 95% prediction interval (PI) of IBM and follow similar behaviour for increasing *x*_*h*_ and *x*_*w*_, although the peak infections for the EBM simulation are higher than the mean of the IBM simulation. These results are aimed at verifying whether the EBM developed here is a good approximation of the IBM.

**Figure 2.**
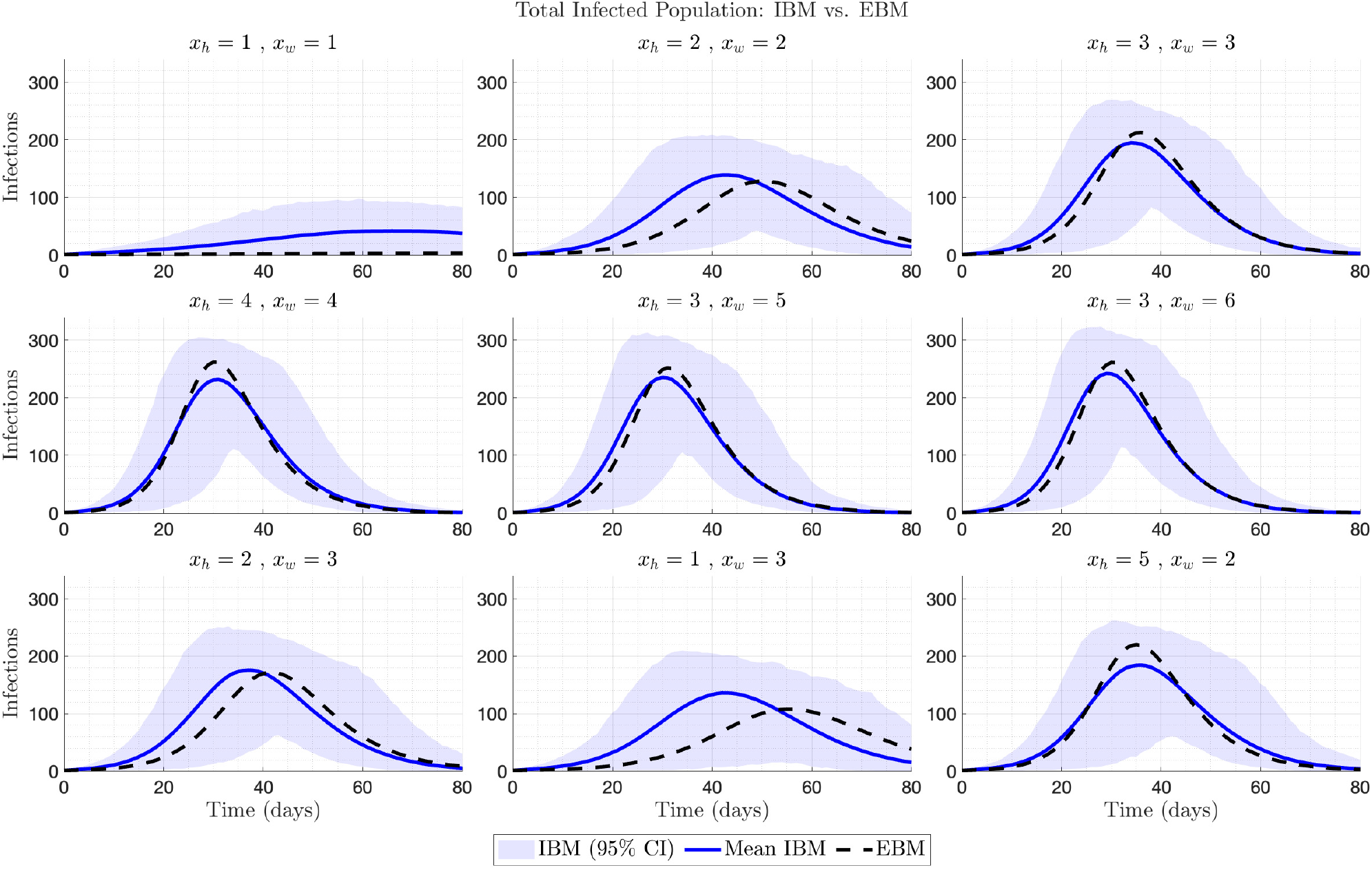
Comparison of total infected population dynamics between the individual-based model (IBM, shaded region showing 95% prediction interval – average of 1,000 realisations) and equation-based model (EBM, solid line) across different external connection configurations. Each subplot represents a specific number of external household (*x*_*h*_) and workplace (*x*_*w*_) connections, demonstrating how increased connectivity affects epidemic progression in both modelling approaches.

**Figure 3.**
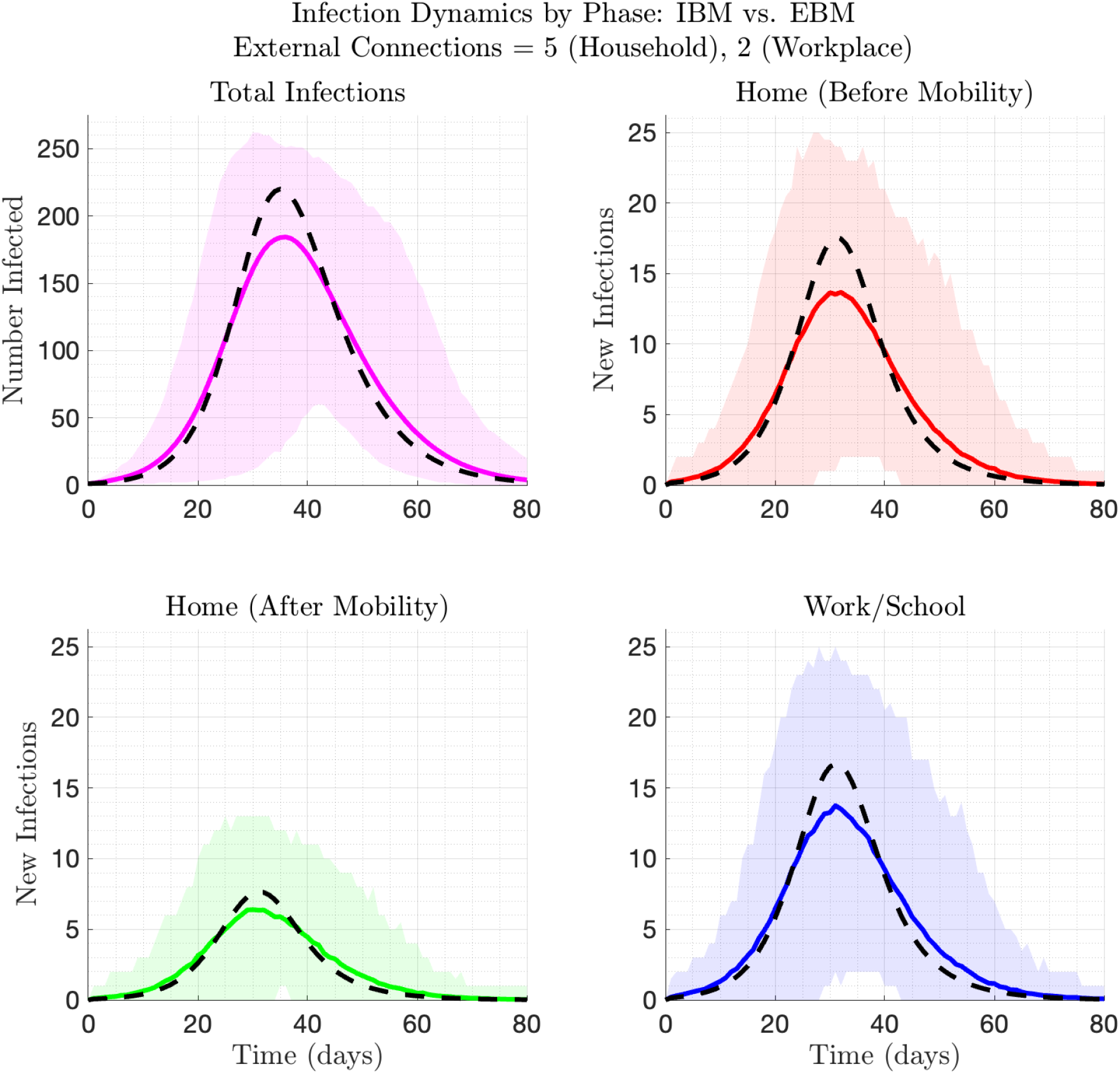
Comparison of transmission phase contributions between IBM (shaded confidence intervals – average of 1,000 realisations) and EBM (solid lines) for configuration with *x*_*h*_ = 5, *x*_*w*_ = 2. (top left) Total infections (top right), home-time household infections (bottom left), work-time household infections (bottom right), and workplace infections.

### 3.1 One at a time (OAT) sensitivity analysis results

The local sensitivity of ℛ_0_ to each parameter, numerically computed from the partial derivatives computed in the methods section, is shown in a tornado plot in Figure 4. The local sensitivity analysis reveals the relative importance of both biological and non-biological parameters on ℛ_0_ (using the baseline parameter values in Table 2). Parameters such as *µ* (biological infectious probability), *c*_*w*_ (average contact per capita per time step), and *α* (mobility probability) exhibit the highest positive sensitivity indices, indicating that increases in these parameters increase ℛ_0_. Conversely, *γ* (the recovery/removal probability) shows a strong negative sensitivity index, suggesting that a higher recovery/removal probability reduces the ℛ_0_. Parameters such as the average external connections at work/school (*x*_*w*_), average household contacts (*c*_*h*_), and the average household external connections (*x*_*h*_) have moderate positive indices, implying a lesser but notable influence on the ℛ_0_. Parameters with the least positive indices are in descending order, the distance threshold required for transmission (*d*), the average work cluster size (*n*_*w*_), the average household size (*n*_*h*_), the average contact duration at work (*T*_*w*_), and at home (*T*_*h*_). Parameters with the least negative indices are the threshold contact duration (*τ*), the average contact proximity at work (*D*_*w*_), and at home (*D*_*h*_).

**Figure 4.**
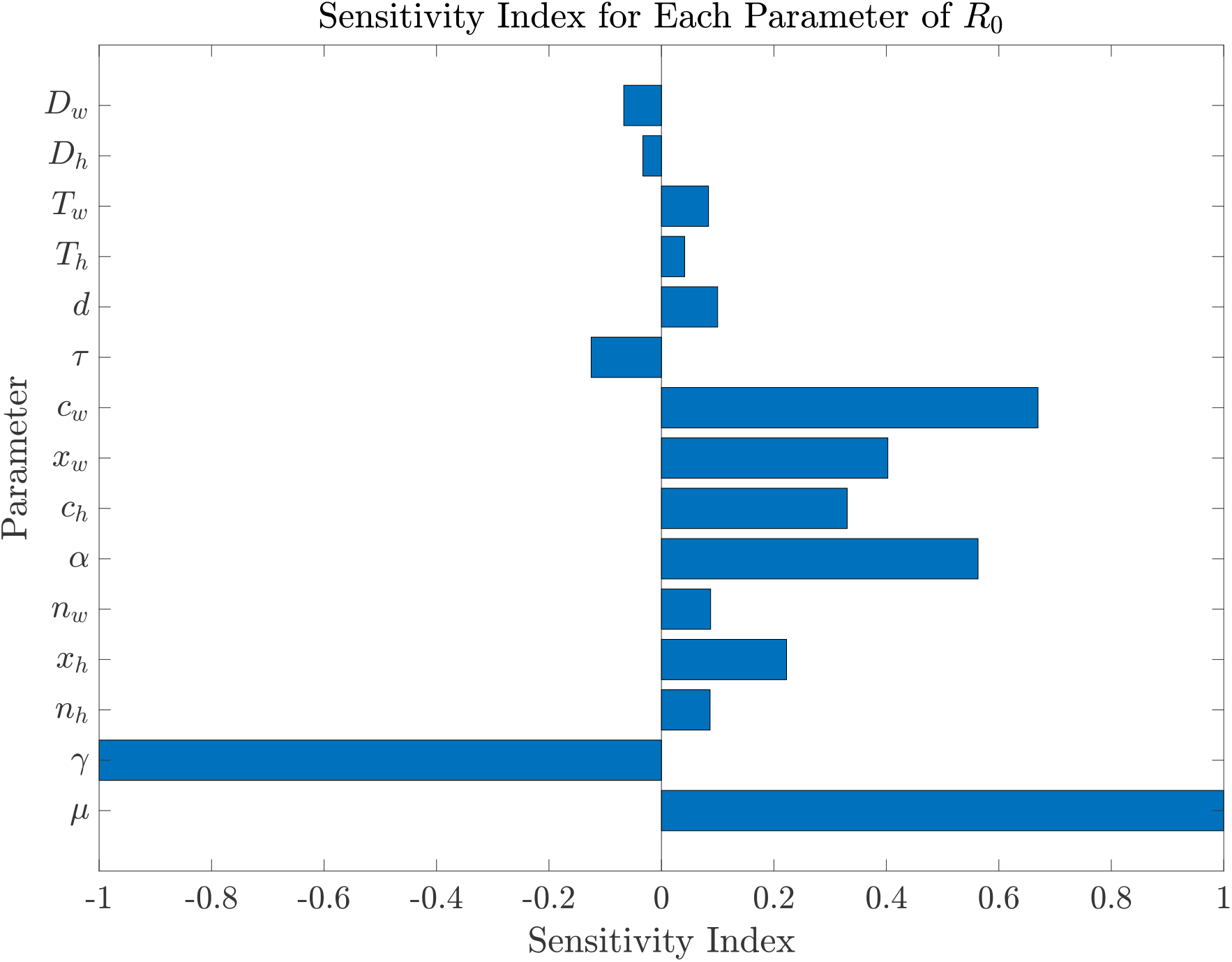
One-at-a-time (OAT) sensitivity indices for the basic reproduction number, ℛ_0_, with respect to model parameters. The sensitivity index *S*_*i*_ measures the relative change in ℛ_0_ due to a small change in parameter *i*, calculated as 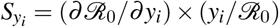. Positive values indicate ℛ_0_ increases with the parameter, while negative values indicate an inverse relationship.

### 3.2 Global sensitivity analysis

The global sensitivity analysis results using the Sobol Sensitivity Analysis Algorithm are presented in Table 3, showing the sensitivity indices and their order of importance, and in Figure 5, which is the visual depiction of the global sensitivity indices. Figure 6 shows how any pair of parameters jointly influences the ℛ_0_. The first-order Sobol indices *S*_*i*_ measure the direct contribution of each parameter to the output variance. The total-order Sobol indices 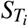 account for both direct effects and interactions between parameters. Here, the parameters with high *S*_*i*_ or 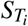 values are more influential on the ℛ_0_.

**Table 3.**
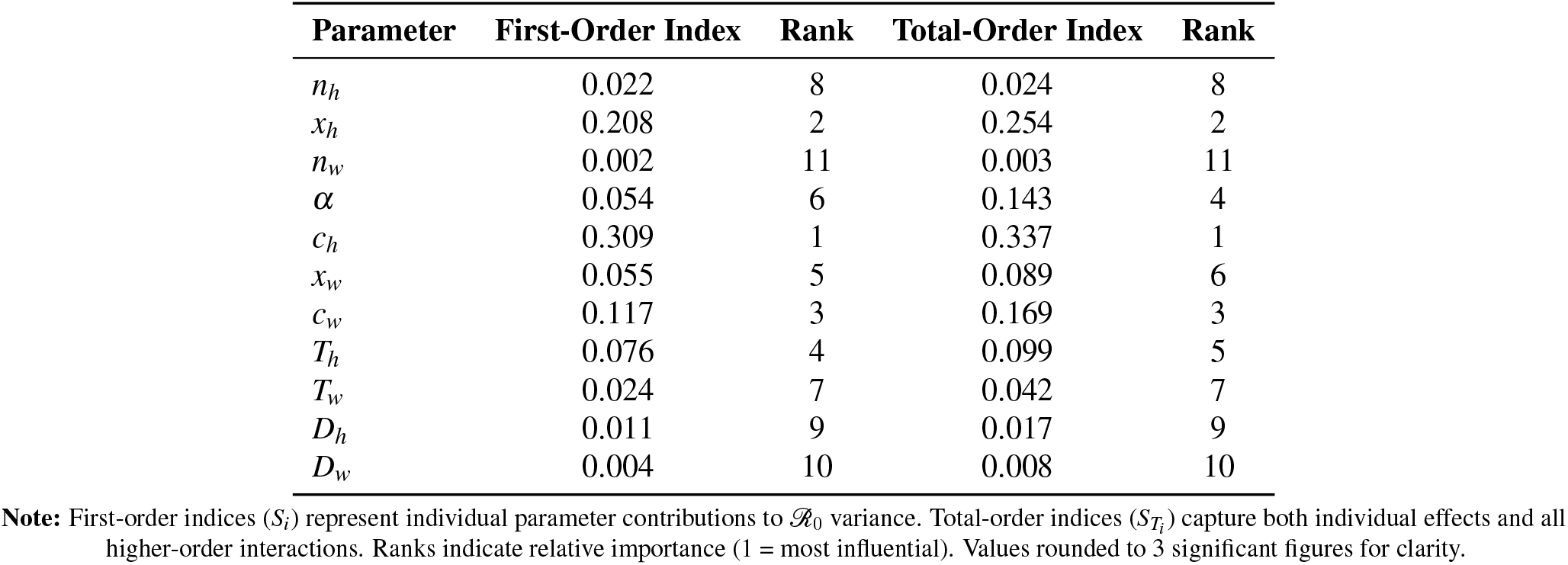
Global sensitivity analysis results for parameters, showing first-order and total-order sensitivities along with their order of importance rankings.

| Parameter | First-Order Index | Rank | Total-Order Index | Rank |
| --- | --- | --- | --- | --- |
| $n_h$ | 0.022 | 8 | 0.024 | 8 |
| $x_h$ | 0.208 | 2 | 0.254 | 2 |
| $n_w$ | 0.002 | 11 | 0.003 | 11 |
| $\alpha$ | 0.054 | 6 | 0.143 | 4 |
| $c_h$ | 0.309 | 1 | 0.337 | 1 |
| $x_w$ | 0.055 | 5 | 0.089 | 6 |
| $c_w$ | 0.117 | 3 | 0.169 | 3 |
| $T_h$ | 0.076 | 4 | 0.099 | 5 |
| $T_w$ | 0.024 | 7 | 0.042 | 7 |
| $D_h$ | 0.011 | 9 | 0.017 | 9 |
| $D_w$ | 0.004 | 10 | 0.008 | 10 |

**Note:** First-order indices ( $S_i$ ) represent individual parameter contributions to $\mathcal{R}_0$ variance. Total-order indices ( $S_{T_i}$ ) capture both individual effects and all higher-order interactions. Ranks indicate relative importance (1 = most influential). Values rounded to 3 significant figures for clarity.

**Figure 5.**
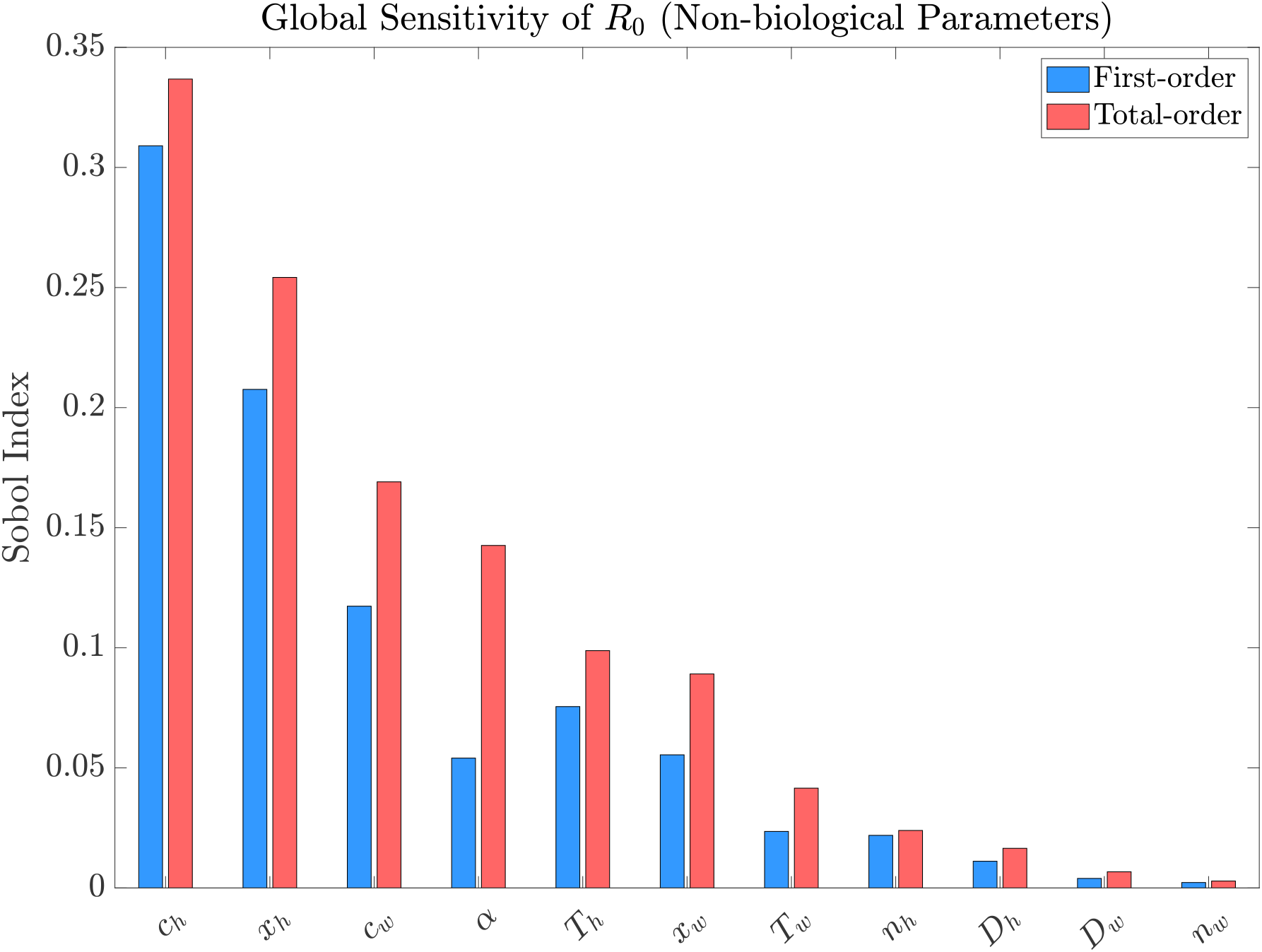
First-order and total-order Sobol sensitivity indices for the basic reproduction number ℛ_0_. Blue bars represent first-order indices (*S*_*i*_), quantifying each parameter’s individual contribution to ℛ_0_ variance. Red bars show total-order indices 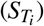, capturing both individual effects and all higher-order interactions.

**Figure 6.**
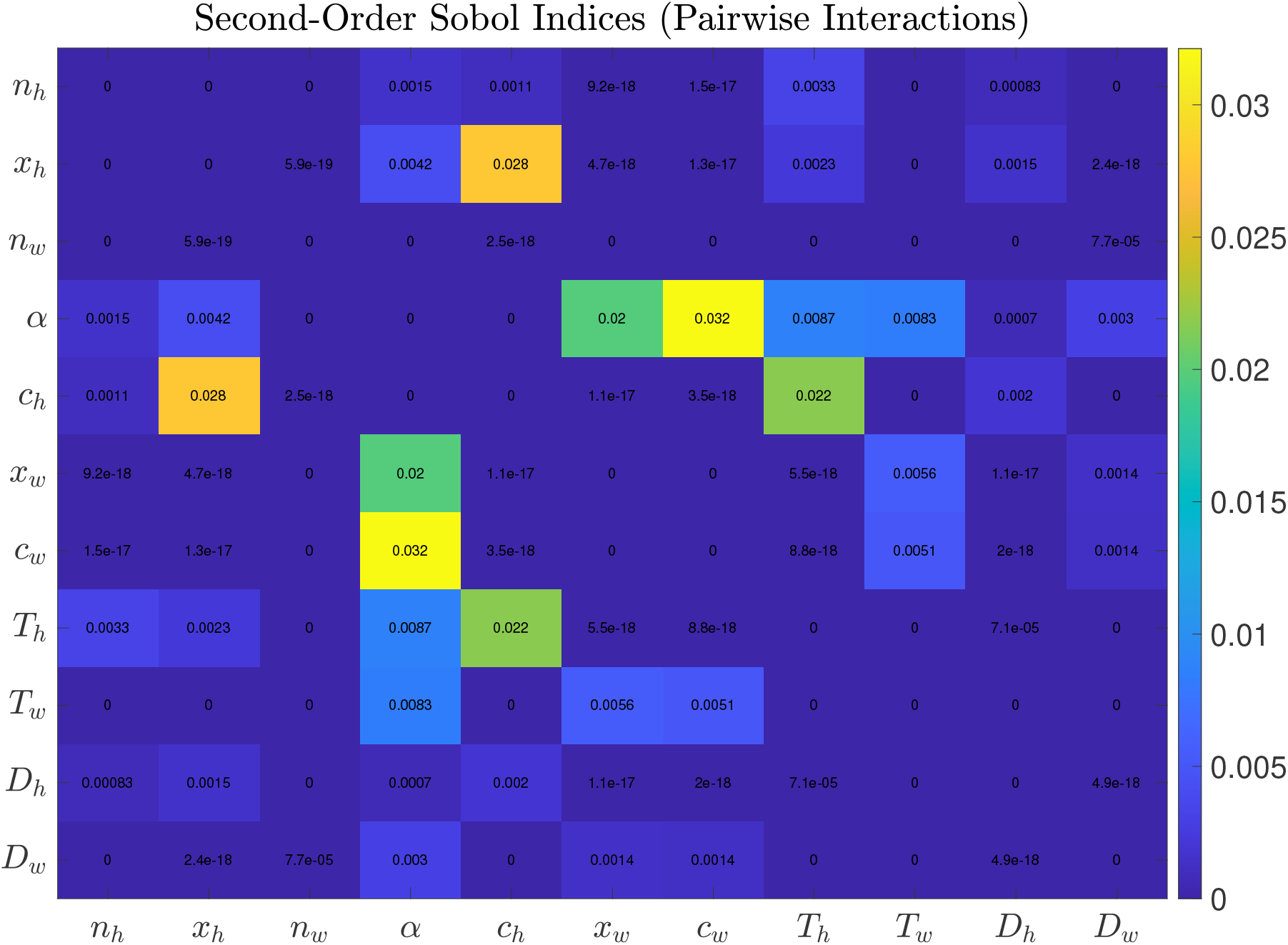
Heatmap of second-order Sobol indices showing pairwise interaction effects between parameters. Each cell represents the joint contribution (*S*_*i j*_) of two parameters to ℛ_0_ variance that cannot be explained by their individual effects alone. Warmer colours indicate stronger interactions, with numerical values showing the proportion of variance explained. The diagonal is blank, as these represent individual effects. The symmetric matrix demonstrates that *S*_*i j*_ = *S* _*ji*_ by definition.

The first-order indices in Figure 5 measure the contribution of a single parameter to the output variance of ℛ_0_, ignoring interactions with other variables. The second-order indices measure the joint effect of two parameters (due to their interaction) that cannot be explained by their individual effects alone, while the total-order indices measure the total contribution of a parameter, including all its individual effects and interactions with other variables. These interactions are analysed across the range of values of the parameters.

The parameters that we can leverage and which have an impact on increasing or decreasing the ℛ_0_ value were chosen, and a 50% change was made in their values OAT while keeping the rest of the parameters as the baseline values. From the results in Figure 4, we increased *D*_*w*_ and *D*_*h*_, while reducing *T*_*w*_, *T*_*h*_, *C*_*w*_, *C*_*h*_, *α, k* and *h* (Figure 7). This is intended to show the variability in the infection dynamics when each parameter value is changed.

**Figure 7.**
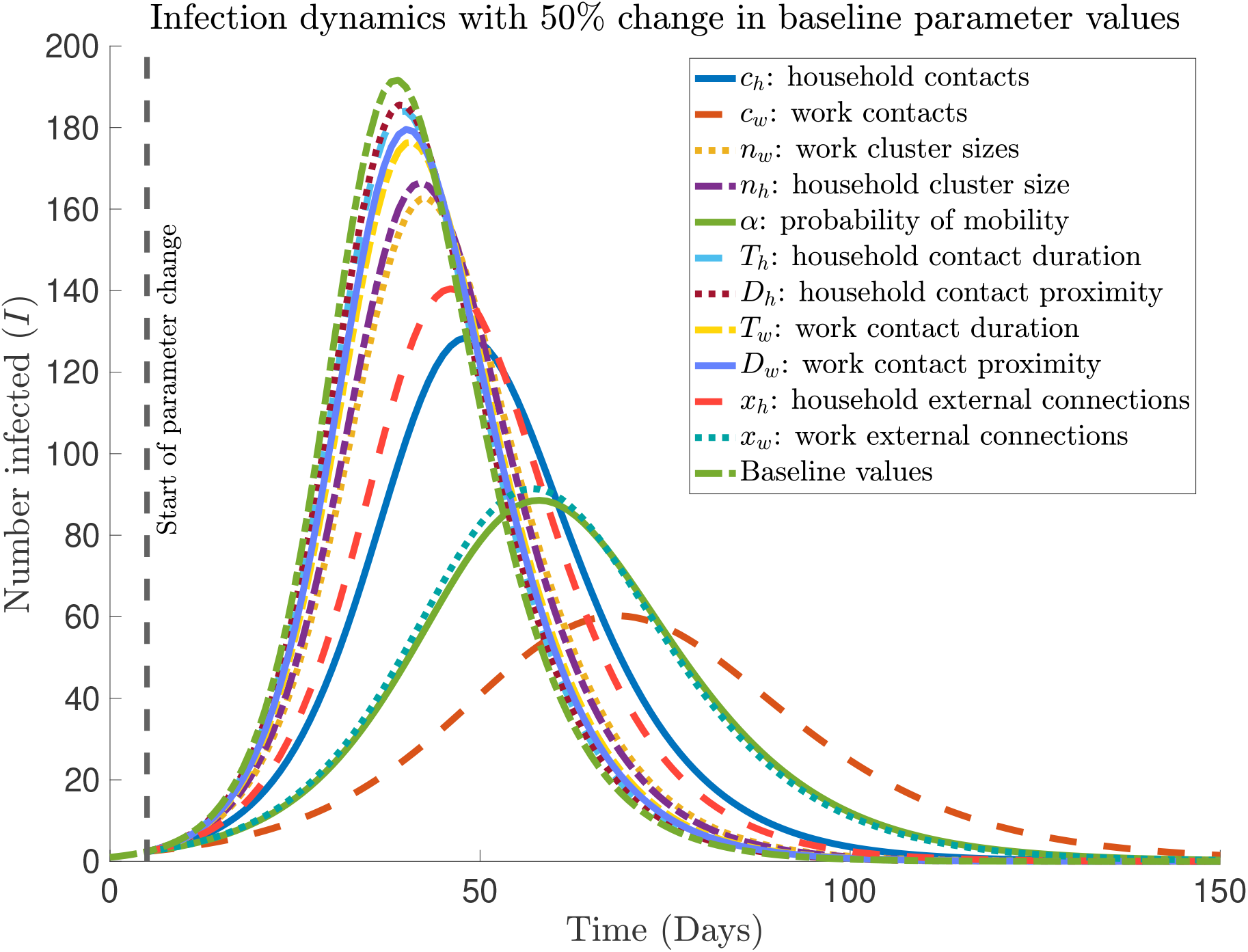
Infection dynamics under varying one-at-a-time (OAT) parameter changes. The plot shows the number of infected individuals (*I*) over time (days) for different OAT parameter perturbations (each with a *±*50% change from baseline values), including contact rates (*c*_*h*_, *c*_*w*_), cluster sizes (*n*_*h*_, *n*_*w*_), mobility probability (*α*), contact durations (*T*_*h*_, *T*_*w*_), contact proximity (*D*_*h*_, *D*_*w*_), and external connections (*x*_*h*_, *x*_*w*_). The black dashed vertical line marks the start of intervention (parameter change) at *t* = 5 days. The baseline SIR model (without parameter changes) is shown for comparison. All simulations use the same initial conditions and model parameters except for the indicated perturbation.

The impact of a 50% change in baseline parameter values in Figure 7 confirms the result of local sensitivity analysis in Figure 4. Similarly to the local sensitivity analysis, the five main parameters that decrease the epidemic peak are *c*_*w*_, *α, x*_*w*_, *c*_*h*_, and *x*_*h*_, reflecting the pivotal role of the number of contacts and external connections within households and work/school, along with mobility. Targeting *n*_*w*_ or *n*_*h*_ (average work and household size) leads to a moderate effect, while changes in *T*_*w*_, *T*_*h*_ (work and household contact duration), *D*_*w*_, and *D*_*h*_ (work and household contact proximity) show a minimal deviation from the baseline result (without change in parameter values).

## 4 Discussion

In this study, we extended our theoretical *Semi-Random Mixing* (SeRaMix) modelling framework [7] to incorporate parameters that explicitly link household and non-household interactions through the mobility of people to and from work to capture the multi-phase interaction in the spread of infection in the population. The approach presented in this work disentangles the parameters representing the social/behavioural contribution from those representing the biological characteristics of the pathogen. It differentiates the various contributions during home-time and work-time interactions occurring within and outside of households, for example, in workplaces. The analysis is aimed at establishing the validity of the proposed EBM and studying the effect of incorporating the multiphased transmission mechanism into a single model, including what the roles of the different parameters mean in the overall dynamics of infectious disease.

Figure 1 shows an increase in the epidemic peak and the early outbreak trajectories with increasing external connections at home (*x*_*h*_) and at work (*x*_*w*_). The result of the IBM simulation confirms the results in [7], which revealed that even when people make the same number of daily contacts, the larger the pool of individuals from which these contacts are randomly chosen (potentially recurrent contact network), the higher the risk of infection in the population and vice versa.

In Figures 2 and 3, the simulation results of the EBM developed in this work are well within a 95% prediction interval of its equivalent IBM. However, the peak infections for the EBM simulation are higher than the mean of the IBM simulation. This phenomenon, where a deterministic model is not strictly the mean of its stochastic counterparts, appears in the literature [7, 32–35], although, for a large population size, a deterministic solution is a good approximation of the stochastic mean [33]. The results show the agreement between IBM and the EBM (Equations (2) - (11)). Thus, this approach is a good method for modelling the spread of infectious diseases in populations with semi-random mixing with both household and non-household interactions.

The notable effects of the transmission probability (*µ*) and the removal/recovery probability (*γ*) ℛ_0_ in Figures 4 and 7 imply that interventions such as testing and isolation of infected people by increasing *γ* will be effective in reducing disease spread. Reducing the number of contacts at work (*c*_*w*_), mobility to work/school through hybrid work (*α*), inter-cluster interaction at work/school (*x*_*h*_), and inter-household interaction (*x*_*h*_) are effective ways to reduce contacts and limit disease spread.

In the global sensitivity analysis, where only non-biological parameters were examined, the effects of parameters *c*_*h*_ (household contacts) and *x*_*h*_ (household external connections) dominate with higher influence (Figure 5), highlighting their individual and interactive effects on ℛ_0_. The total-order indices, slightly higher than first-order indices, suggest strong pairwise interactions, particularly for *c*_*h*_ and *x*_*h*_. This indicates that household contacts and external connections not only affect ℛ_0_ independently but also amplify each other’s impact, necessitating targeted interventions in household settings. Since reducing household contacts may not be feasible, the available option is to reduce the inter-household interaction (*x*_*h*_).

The second-order indices matrix (Figure 6) reveals strong pairwise interactions, with notable values such as *c*_*w*_ and *α* (0.32), *c*_*h*_ and *x*_*h*_ (0.028), and *T*_*h*_ and *c*_*h*_ (0.022). These high interaction indices suggest that work contacts and work mobility (*c*_*w*_ and *α*), household contacts and household external connections (*c*_*h*_ and *x*_*h*_), and contact duration at home and household contacts (*T*_*h*_ and *c*_*h*_) create synergistic effects on the ℛ_0_. Such interactions imply that reducing these parameters jointly could have compound benefits.

The discrepancy in the order of importance of parameters in Figures 4 and 7, which used the baseline parameters used for this study, and the results in Figures 5 and 6 which allow plausible ranges of parameters to be studied, indicate that results of epidemic models are context dependent and may not be universally applicable to all populations and/or pathogens. These insights show that care needs to be taken not to generalise the results of a particular model to all populations or pathogens, particularly when predicting the cause of an epidemic and designing public health control strategies. Although the order of importance varies, both local and global sensitivity analyses indicate that work/school contacts (*c*_*w*_), work/school-based mobility (*α*), household contacts (*c*_*h*_), and household external connections (*x*_*h*_) are the key drivers of the spread of infectious diseases.

The contact size model proposed here provides a flexible framework for incorporating disease-specific thresholds of contact duration and proximity into epidemiological models. It extends prior work by employing modified sigmoid functions to define a bounded, multiplicative *effective contact size* (*C*) that represents how transmission risk increases smoothly with longer and closer interactions. Empirical contact studies, such as the POLYMOD survey [36], and network-based analyses [37], have demonstrated that the duration and proximity of interactions are key determinants of transmission risk. The proposed formulation introduces tunable transitions around pathogen-specific limits, with steepness parameters (*ν*_*τ*_ and *ν*_*d*_) that can be calibrated to empirical data. Unlike computationally intensive agent-based or temporal network models that capture detailed individual heterogeneity [37], this approach aggregates to population averages, making it readily integrable into compartmental frameworks such as SIR/SEIR, where *C* modulates the transmission rate. While commuter and mobility-based metapopulation models emphasise large-scale movement flows between regions [38, 39], this contact-centric framework focuses on local mixing intensity within populations. Moreover, it can complement such models by refining within-patch transmission according to setting-specific contact characteristics—such as the high proximity and short duration typical of public transport versus more distanced, longer interactions in office environments—thereby linking micro-scale behavioural dynamics with macro-scale spatial epidemiology.

Several limitations should be acknowledged. First, the formulation of the contact size function (1) has not been parametrised by any real-world data and therefore is only an illustration of our assumed effect of contact duration and proximity. This study is an attempt to stimulate further research on how to mathematically incorporate these metrics into an epidemic model and how this could help in the design and analysis of non-pharmaceutical interventions. Second, the exclusion of biological parameters (*µ, γ, d, τ*) from the global sensitivity analysis, second-order Sobol indices, and infection dynamics, while practical, limits the study’s ability to assess the full spectrum of factors influencing the basic reproduction number ℛ_0_. This omission may underestimate the combined effects of biological and non-biological parameters, particularly in scenarios where pathogen characteristics vary or where pharmaceutical interventions or NPIs could directly impact factors such as transmissibility/susceptibility or duration of infectious period. Third, the coupling parameter *κ* was as fitted to IBM simulations and used in [7] for a single population size (*N* = 1, 000) and pathogen infectivity and recovery probabilities (*µ* = 0.18, *γ* = 0.16); its generalisation across different pathogens with varying infectious and recovery probabilities and varying population sizes needs further study. Finally, the study assumes uniform household and cluster sizes and external connections, which do not reflect real-world variability. Future research could integrate heterogeneous populations and biological parameters in the global sensitivity analysis, test dynamic assumptions with empirical data, and explore a wider range of parameter values to add robustness.

## 5 Conclusion

We developed a multi-phase transmission model to explicitly incorporate household and non-household interactions into a single equation-based epidemic model with characteristic semi-random mixing behaviour. Three infection phases have been incorporated into the model: i) home-time household infection, where the force of infection takes account of all infected individuals in the population (assuming that everyone returns to their household after work/school), ii) work-time household infection, where the force of infection takes account of all infected individuals who have not moved to work/school from their households, and iii) work-time work/school infections, where the force of infection is computed using only the proportion of people who have moved to work/school per unit of time. A sigmoid-like function is proposed to incorporate the effect of contact duration and proximity between contacts into equation-based models. The basic reproduction number ℛ_0_ is derived analytically, and sensitivity analyses have been used to investigate the influence of different parameters on the reproductive number and infection dynamics.

Using the basic reproduction number, numerical simulation, and both local and global sensitivity analysis, we have demonstrated that this equation-based model framework agrees with its individual-based model counterpart and can be applied in epidemic modelling to add to the understanding of the effects of different parameters for household and non-household interactions on infectious disease spread.

## Data Availability

All data produced in the present study are available upon reasonable request to the authors

## Acknowledgements

This research is fully funded by the Institute for Global Pandemic Planning, Warwick Medical School, University of Warwick, UK.

## Author contributions statement

Conceptualisation: M. L. S.

Methodology: M. L. S.

Supervision: A.S. & K. S. R.

Software: K. S. R. & M. L. S.

Visualisation: K S. R & M. L. S.

Writing – original draft: M. L. S.

Writing – review & editing: A. S., K. S. R. & M. L. S.

## Data availability

MATLAB code will be accessible via GitHub upon publication of the manuscript.

## Ethics declarations

This study focuses on the theoretical development of a mathematical model for the spread of a hypothetical infectious disease. No data was collected and analysed.

