## Supplementary Information for "Semi-Random Mixing Epidemic Model: Integrating Explicit Household and Non-household Interactions"

### ABSTRACT

Supplementary Information

### 1 Modelling contact duration and proximity

Here, we propose the use of a modified sigmoid function to account for the impacts of contact duration and proximity between contacts in models of the effective contact size. Suppose that the threshold contact duration required for an infection to be transmitted is given by $\tau$, and the threshold distance between contacts to transmit infection is $d$. If the average contact duration in the population is $T$, and the average distance between contacts is $D$. Then we assume that the risk of transmission increases with the increase in *Effective contact size* defined by:

$$C = cf(T;\tau)g(D;d), \quad D \geq 0, T > 0, \tag{1}$$

where $f(T;\tau) = \frac{2}{1+e^{\nu_\tau(\tau-T)}}$, is the Duration Risk index represents the effect of contact duration on the transmission and $g(D;d) = \frac{2}{1+e^{-\nu_d(d-D)}}$, is the Proximity Risk index representing the effect of contact proximity on the transmission, $c \geq 1$ is the average number of contacts per unit of time, $\nu_\tau$ is the steepness parameter that represent the speed of effective contact duration, and $\nu_d$ is the steepness parameter representing the speed of effective contact distance. The steepness parameters control how sharply the functions ($f(T;\tau)$ and $g(D;d)$) transition from low to high around the midpoint. As T increases, the chance of transmission increases, and as D increases, the chance of transmission decreases.

The functions $f(T;\tau)$ and $g(D;d)$ have been constructed by modifying the sigmoid function [1, 2] such that a transition exists around the threshold required for an infection to be transmitted through person-to-person contact.

The analysis of the behaviour of the function $f(T;\tau)$ is as follows:

- When $\tau > T$:

  In this case, $\tau - T > 0$, leading to $1 + e^{\nu_\tau(\tau-T)}$ tending to infinity, and $f(T;\tau)$ approaches 0.

  This indicates a reduced impact of contact (on transmission) when the average contact duration is less than the threshold duration required for transmission.

- When $\tau = T$:

  This case is the threshold condition which implies $\tau - T = 0$, where $e^{\nu_\tau(\tau-T)}$ becomes 1, leading to $f(T;\tau)$ = 1

  This satisfies the threshold condition where the risk of transmission is determined by the number of contacts when the average contact duration equals the threshold required duration.

- When $\tau < T$:

In this case, $\tau - T < 0$, leading to $e^{\nu_\tau(\tau - T)}$ decreasing towards 0, and $1 + e^{\nu_\tau(\tau - T)}$ approaches 1, and $f(T;\tau)$ approaches 2.

This indicates an increased risk of contact (on transmission) when the average contact duration exceeds the threshold required duration to transmit infection. However, this value asymptotes to 2. This result is reasonable, as although an increase in contact duration increases the risk of transmission, this risk does not increase indefinitely.

Equation (1) (depicted in Fig. 1) ensures that the shorter the distance between contacts, the higher the contact size, and the longer the average contact duration, the higher the contact size, however, these quantities are interdependent such that although a person may have a longer duration of contacts, such contacts may not be within proximity and therefore may not increase the contact size, and vice versa. Hence, it is important to consider the region around the contact size threshold where the spread of diseases could be amplified (see Fig. 2). With the concept of contact size, we can analyse the effect of social distancing by targeting and increasing $D$. We can gain insight into the effect of contact duration by targeting $T$.

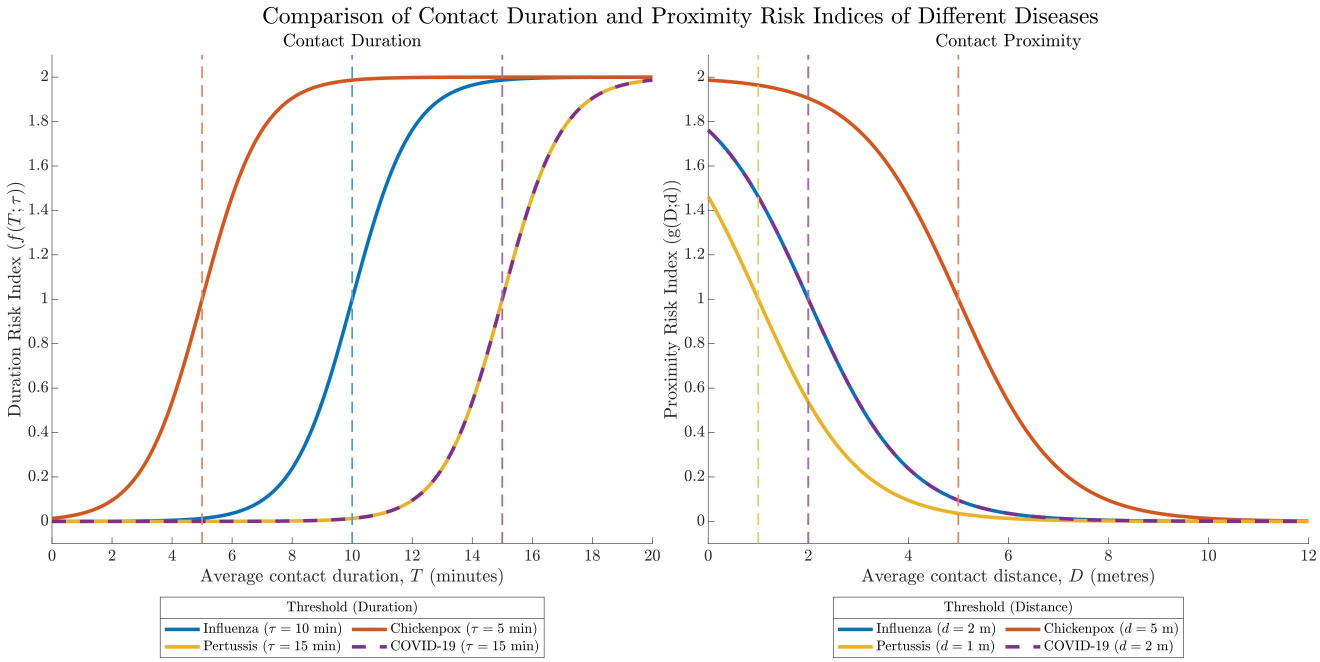


**Figure 1.** Risk indices for disease transmission as functions of contact duration and proximity. (Left) Duration risk showing sigmoidal dependence on average contact time $T$ relative to disease-specific thresholds $\tau$. (Right) Proximity risk demonstrating dependence on average contact distance $D$ relative to thresholds $d$. Dashed vertical lines indicate critical thresholds for each disease: COVID-19 ($\tau = 15$ min, $d = 2$ m), Influenza ($\tau = 10$ min, $d = 2$ m), Chickenpox ($\tau = 5$ min, $d = 5$ m), and Pertussis ($\tau = 15$ min, $d = 1$ m). Risk scales from 0 (no risk) to 2 (maximum risk), with the transition midpoint at each disease's threshold value. In all cases, $c = \nu_\tau = \nu_d = 1$.

#### 1.1 Modelling household and non-household interactions

The SeRaMix per-contact probability of meeting an infected person in the population at time $t$ in [3] was given by:

$$p(t) = \frac{I(t)}{N}\left(1 - e^{-\kappa \frac{nx}{n+1}}\right), \qquad (2)$$

where $I(t)$ is the total number of infected individuals at time $t$, $N$ is the total population (irrespective of infection status), $n$ is the average size of the cluster in the population, $x$ is the average number of external connections per capita, and $\kappa$ is a constant that indicates the connectivity of the clusters in the population. Although this modified force of infection does not explicitly specify whether the clusters are defined as household groups or work/school groups, these distinctions exist in real life, and it is clear that an average person has (potentially) recurrent contacts within the household and another set at work/school. Thus, typically, a person will make a random choice of contacts from all recurrent contacts at two levels each day: at home and at work/school.

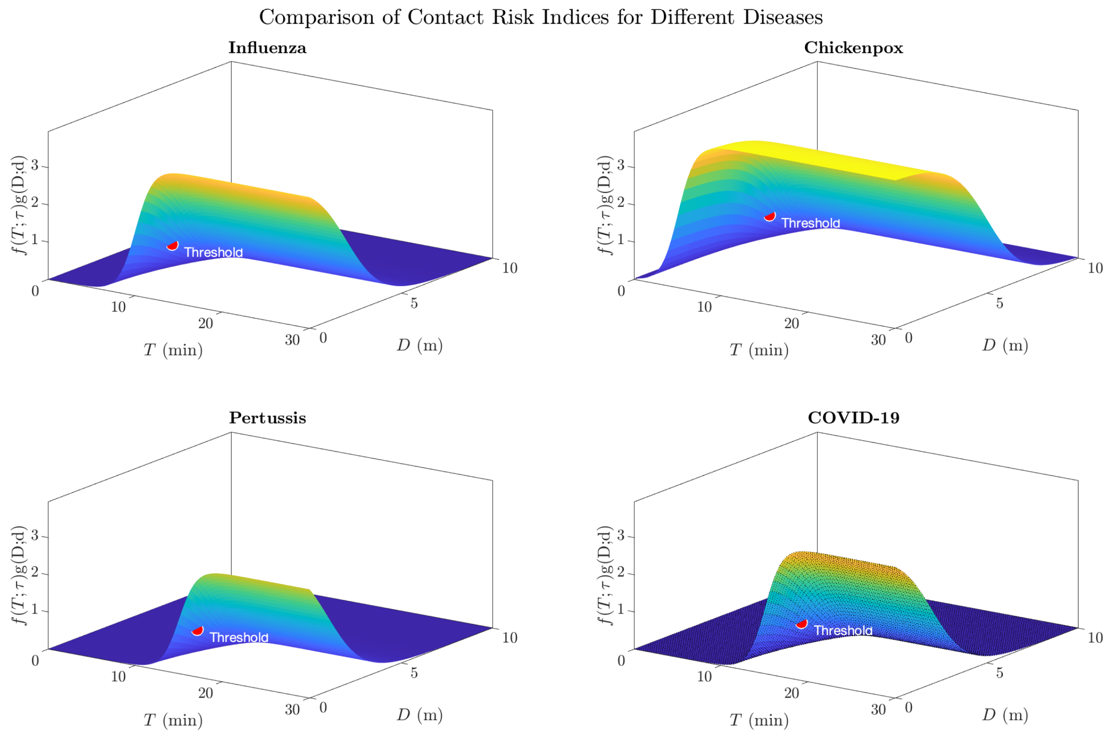


**Figure 2.** Three-dimensional visualisation illustrating contact risk indices as a function of contact duration ($T$) and distance ($D$) for four example infectious diseases. Each subplot shows the sigmoidal contact risk index, where $\tau$ represents the disease-specific duration threshold and $d$ the distance threshold. The red markers indicate the threshold points where $T = \tau$ and $D = d$. All surfaces share the same colour scale for risk comparison. Parameters used: COVID-19 ($\tau = 15$ min, $d = 2$ m), Influenza ($\tau = 10$ min, $d = 1.5$ m), Chickenpox ($\tau = 5$ min, $d = 1$ m), and Pertussis ($\tau = 20$ min, $d = 0.5$ m). In all cases, $c = \nu_\tau = \nu_d = 1$.

To formulate the multi-stage interactions and infection, we use the steps in **Algorithm Box 1**. First, we consider that at each time step, a proportion $\alpha \geq 0$ of people will leave their homes for work/school, while a proportion $(1-\alpha)$ will remain home, where non-work-related interaction may occur. Let $S(t)$, $I(t)$, $R(t)$ be the numbers of the susceptible, infected, and recovered (removed) individuals at time $t$, respectively. We assume that all individuals return home after work-time interactions and are at risk of being infected at home during the home-time interaction through the household probability of infection. Furthermore, we assume that during work-time interactions, any susceptible person who does not move to work remains and interacts with other people at home and can only be infected by infected persons who do not go to work. Susceptible individuals who go to work have the risk of being infected during work-time interaction outside their households. We assume that infections that occur within household areas after the return stage and before the next movement stage are counted as infections of the next time step occurring during home-time interaction. Thus, the daily multi-level infection begins with the home-time infection before the movement stage and ends when people return home.

Let the home-time household force of infection (FOI) (the probability that a susceptible person will be infected at home during home-time interaction) be denoted by $P_h(t)$, work-time household FOI (the probability that a susceptible person will be infected at home during work-time interaction) as $P_{hw}(t)$, and non-household work-time FOI (the probability that a susceptible person will be infected outside the home during work-time interaction) as $P_w(t)$. Here, the expected number of new infections resulting from home-time interactions would be $S(t)P_h(t)$. The population $S(t) - S(t)P_h(t)$ that remains susceptible after the home-time interaction can be infected at home or workplaces/schools during the work-time interaction. Combining these, we have the new infections expected in the next time step:

$$S(t)P_h(t) + S(t)\Big(1 - P_h(t)\Big)\Big((1-\alpha)P_{hw}(t) + \alpha P_w(t)\Big), \quad (3)$$

**Algorithm 1** Discrete-Time Multi-Stage Interaction Algorithm

**Input:** Population $N$, mobility proportion $\alpha$, infection rates $\beta_h$, $\beta_w$, recovery probability $\gamma$, cluster sizes $n_h$ (home), $n_w$ (work), external connections $x_h$ (home), $x_w$ (work), coupling parameter $\kappa$, time steps *MaxTime*.
**Output:** Counts of susceptible ($S$), infected ($I$), recovered ($R$) per time step.
**Initialise:** initialise arrays for $S$, $I$, $R$.
**for** each time step $t = 0$ to *MaxTime* **do**
    **Home Interaction:**
    **for** all susceptible in households **do**
        Compute the force of infection involving all infected individuals (assuming all return home after work) $P_h(t)$
        Compute new infections within households using force of infection $P_h(t)$.
    **end for**
    **Mobility:**
    Select proportion $\alpha$ of population to move to work.
    **Work-Time Household Interaction:**
    **for** all susceptible remaining in households **do**
        Compute the force of infection involving all infected non-mobile individuals $P_{hw}(t)$
        Compute new infections within households using force of infection $P_{hw}(t)$ for non-mobile individuals (proportion $1-\alpha$).
    **end for**
    **Work Interaction:**
    **for** all susceptible at work **do**
        Compute the force of infection involving all infected mobile individuals
        Compute new infections at work using force of infection $P_w(t)$ for mobile individuals (proportion $\alpha$).
    **end for**
    **Recovery:**
    **for** all infected individuals from the previous time step **do**
        Compute number of recoveries.
    **end for**
    **Record:** Store $S$, $I$, $R$ for time $t$.
**end for**
$\triangleright$ Day ends

which simplifies to:

$$S(t)\left[P_h(t)+\left(1-P_h(t)\right)\left((1-\alpha)P_{hw}(t)+\alpha P_w(t)\right)\right]. \tag{4}$$

The household and non-household force of infection terms are dependent on the cluster size in the different places. We assume the average home cluster size is $n_h$ during the household-time interactions, and this reduces to $(1-\alpha)n_h$ after mobility due to a proportion $\alpha$ of the residence not being present during this time. We assume the average cluster size at work for the mobile individuals is $\alpha n_w$. We also assume that $x_h$ is the average number of external connections per person at home and $x_w$ is the average number of external connections per person at work/school. This is to incorporate the effect of mobility on the cluster sizes.

The force of infection terms are then given by the following equations (2):

$$P_h(t)=\beta_h\frac{I(t)}{N}\left(1-e^{-\kappa\frac{n_h x_h}{n_h+1}}\right), \tag{5}$$

$$P_{hw}(t)=\beta_h\frac{(1-\alpha)I(t)}{(1-\alpha)N}\left(1-e^{-\kappa\frac{(1-\alpha)n_h x_h}{(1-\alpha)n_h+1}}\right)=\beta_h\frac{I(t)}{N}\left(1-e^{-\kappa\frac{(1-\alpha)n_h x_h}{(1-\alpha)n_h+1}}\right), \tag{6}$$

$$P_w(t)=\beta_w\frac{\alpha I(t)}{\alpha N}\left(1-e^{-\kappa\frac{\alpha n_w x_w}{\alpha n_w+1}}\right)=\beta_w\frac{I(t)}{N}\left(1-e^{-\kappa\frac{\alpha n_w x_w}{\alpha n_w+1}}\right). \tag{7}$$

In addition, $\beta_h=\mu C_h$, where $\mu$ is the pathogen-dependent probability that infection will be transmitted after contact with an infected person, and $C_h$ is the average household contact size, and $\beta_w=\mu C_w$, where $C_w$ is the average work contact size.

The household *effective contact size* is given by:

$$C_h=c_h f(\tau,T_h)g(d,D_h), \qquad D_h\geq 0, T_h>0, \tag{8}$$

where $c_h$ is the average household contacts, $T_h$ is the average household contact duration, and $D_h$ is the average household contact proximity, and the work *effective contact size* is given by:

$$C_w=c_w f(\tau,T_w)g(d,D_w), \qquad D_w\geq 0, T_w>0, \tag{9}$$

where $c_w$ is the average work contact, $T_w$ is the average contact duration, and $D_w$ is the average contact distance outside the residence/household.

The total FOI on day $t$ is thus given by:

$$\Delta(t)=P_h(t)+\left(1-P_h(t)\right)\left((1-\alpha)P_{hw}(t)+\alpha P_w(t)\right). \tag{10}$$

Using a similar approach in [3], we can compute the $P_h(t)$, $P_{hw}(t)$, and $P_w(t)$ as:

$$P_h(t)=1-\left(1-\mu p_h(t)\right)^{C_h}. \tag{11}$$

$$P_{hw}(t)=1-\left(1-\mu p_{hw}(t)\right)^{C_h}. \tag{12}$$

$$P_w(t)=1-\left(1-\mu p_w(t)\right)^{C_w}. \tag{13}$$

The terms $p_h(t)$, $p_{hw}(t)$, and $p_w(t)$ as adapted from equation (2) are given by:

$$p_h(t)=\frac{I(t)}{N}\left(1-e^{-\kappa\frac{n_h x_h}{n_h+1}}\right), \tag{14}$$

$$p_{hw}(t)=\frac{I(t)}{N}\left(1-e^{-\kappa\frac{(1-\alpha)n_h x_h}{(1-\alpha)n_h+1}}\right), \tag{15}$$

$$p_w(t)=\frac{I(t)}{N}\left(1-e^{-\kappa\frac{\alpha n_w x_w}{\alpha n_w+1}}\right). \tag{16}$$

### 1.2 Model validation and analysis

An IBM approach can provide granular insight into epidemic spread, complementing equation-based models (EBMs) by capturing heterogeneous mixing patterns, small-scale clustering effects, and stochastic variability in early outbreaks. The framework enables systematic investigation of how household/workplace sizes ($n_h$, $n_w$), a measure of mobility ($\alpha$), contact rates ($c_h$, $c_w$) and external connectivity ($x_h$, $x_w$) collectively influence epidemic trajectories and transmission patterns.

Similar to the approach in [3], our analogous IBM (**Algorithms** 2–6) is used here to validate the ability of the EBM developed in this study.

**Algorithm 2** Function to Construct Contact Network (Part A)

```
Input: Total population N, cluster size n_h/n_w, number of external connections num_ext
Output: Neighbour matrix with N rows and n_h/n_w − 1 + num_ext columns
Assign Clusters:
Randomly shuffle all N individuals
Group individuals into clusters of size n_h/n_w
Set Up Storage:
Create empty lists for each agent's cluster neighbours
Create empty lists for each agent's external neighbours
Create neighbour matrix of size N × (n_h/n_w − 1 + num_ext) filled with placeholder values
Add Cluster Neighbours:
for each cluster do
    Get the list of individuals in the cluster
    for each agent in the cluster do
        Add all other cluster individuals (except the agent itself) to their cluster neighbour list
    end for
end for
Add External Neighbours:
if num_ext > 0 then
    Set each agent to need num_ext external neighbours
    while some individuals need more external neighbours do
        Find individuals who still need external neighbours
        if 2 to 4 individuals remain then
            for each agent in sequence (except the last) do
                Pair current agent with the next agent
                if both individuals need more external neighbours then
                    Add each agent to the other's external neighbour list
                    Reduce each agent's needed connections by 1
                end if
            end for
            Stop adding external neighbours
        end if
        if fewer than 2 individuals remain then
            Stop adding external neighbours
        end if
    end while
end if
```

Except where indicated, the baseline parameter values in Table 1 have been used for simulation.

### 1.3 Derivation of the Reproduction Number

We derived the basic reproduction number $\mathscr{R}_0$ using the next generation matrix (NGM) approach for the discrete-time model [4, 5]:

$$\mathscr{R}_0 = \rho\left(F[\mathbb{I} - T]^{-1}\right), \tag{17}$$

where $\rho$ is the spectral radius, $F$ is the transmission matrix, $\mathbb{I}$ is the identity matrix, and $T$ is the transition matrix for the infected compartment $I(t)$.

**Algorithm 3** Function to Construct Contact Network (Part B)

Randomly shuffle remaining individuals
**for** each agent in the shuffled list **do**
    Get current agent
    Exclude the agent, their cluster neighbours, and existing external neighbours
    Find possible new neighbours who still need connections
    **if** no possible neighbours exist **then**
        Skip to the next agent
    **end if**
**end for**
Randomly pick one possible neighbour
**if** both individuals need more external neighbours **then**
    Add each agent to the other's external neighbour list
    Reduce each agent's needed connections by 1
    **if** both individuals have all the needed connections **then**
        Remove both individuals from the list
    **end if**
**end if**
**if** all individuals have all needed connections **then**
    Stop adding external neighbours
**end if**
**Build Neighbour Matrix:**
**for** each agent **do**
    Combine their cluster and external neighbour lists
    **if** combined list is shorter than $n_h - 1 + \texttt{num\_ext}$ **then**
        Add zeros to fill the list to length $n_h - 1 + \texttt{num\_ext}$
    **end if**
    Set the agent's row in the neighbour matrix to the combined list
**end for**
**Return:** Neighbour matrix

**Algorithm 4** SIR Semi-Random Mixing Individual-Based Model with Household and Workplace Clustering (Part A)

**Input:** Structure `para` with $N$, $\mu$, $\gamma$, `MaxTime`, `Realisations`, `OutbreakThreshold`, `household_size` ($n_h$), `cluster_size` ($n_w$), $\alpha$, $c_h$, $c_w$, `extenH` ($x_h$), `extenW` ($x_w$)
**Output:** Counts of susceptible ($S$), infected ($I$), removed ($R$), and infections by source (`HNew`, `HWNew`, `WNew`)
**Initialisation:**
Initialise arrays for $S$, $I$, $R$, `HNew`, `HWNew`, `WNew` infections
Initialise cells for confidence intervals
**for** each configuration $idx = 1$ to length$(x_h)$ **do**
    Set $x_h[idx], x_w[idx]$ as external household/workplace connections
    **Network Construction:**
    `household_neighbours` $\leftarrow$ build_network$(N, n_h, x_h[idx])$ ▷ Household contacts
    `work_neighbours` $\leftarrow$ build_network$(N, n_w, x_w[idx])$ ▷ Workplace contacts
    **Simulate Realisations:**
    **while** successful realisations $<$ `Realisations` **do**
        Initialise `health_matrix` ($S = 0, I = 1, R = 2$)
        Randomly select one agent and set it as infected
        Set $S \leftarrow N-1, I \leftarrow 1, R \leftarrow 0$
        **for** each time step $t = 2$ to `MaxTime` $+ 1$ **do**
            Copy previous health states
            Initialise infection counters
            **Phase 1: Home Infection (Before Mobility):**
            **for** each susceptible agent $i$ **do**
                **Contact Selection:**
                Randomly select min$(c_h, |\text{household neighbours}|)$ contacts
                Count infected contacts $k$ from selected contacts
                **Infection Dynamics:**
                Generate random number $r \in [0, 1]$
                **if** $r < \mu \cdot k$ **then**
                    Set agent $i$ as infected
                    Increment `HNew`
                **end if**
            **end for**
        **end for**
    **end while**
**end for**

**Algorithm 5** SIR Semi-Random Mixing Individual-Based Model with Household and Workplace Clustering (Part B)

**Mobility Decision:**
Set `is_mobile` for each agent with probability $\alpha$
**Phase 2: Home Infection (After Mobility):**
**for** each susceptible, non-mobile agent $i$ **do**
  **if** $c_h \leq |\text{home neighbours}|$ **then**
    **Contact Selection:**
    Randomly select $c_h$ contacts from non-mobile household neighbours
    Count infected contacts $k$ from selected contacts
    **Infection Dynamics:**
    Generate random number $r \in [0,1]$
    **if** $r < \mu \cdot k$ **then**
      Set agent $i$ as infected
      Increment `HWNew`
    **end if**
  **end if**
**end for**
**Phase 3: Workplace Infection:**
**for** each agent $i$ **do**
  **if** agent $i$ is susceptible and `is_mobile`[$i$] **then**
    **if** $c_w \leq |\text{work neighbours}|$ **then**
      **Contact Selection:**
      Randomly select $c_w$ contacts from mobile work neighbours
      Count infected contacts $k$ from selected contacts
      **Infection Dynamics:**
      Generate random number $r \in [0,1]$
      **if** $r < \mu \cdot k$ **then**
        Set agent $i$ as infected
        Increment `WNew`
      **end if**
    **end if**
  **else if** agent $i$ is infected **then**
    **Recovery Dynamics:**
    Generate random number $r \in [0,1]$
    **if** $r < \gamma$ **then**
      Set agent $i$ as removed
    **end if**
  **end if**
**end for**

**Algorithm 6** SIR Semi-Random Mixing Individual-Based Model with Household and Workplace Clustering (Part C)

**for** each configuration $idx = 1$ to length$(x_h)$ **do**
  **while** successful realisations $<$ `Realisations` **do**
    **for** each agent $i$ **do**
      **Record State:**
      Update counts: $S \leftarrow \sum(\text{susceptible})$, $I \leftarrow \sum(\text{infected})$, $R \leftarrow \sum(\text{removed})$
    **end for**
    **Check Outbreak:**
    **if** $R_{\text{final}} \geq$ `OutbreakThreshold` **then**
      Store $S$, $I$, $R$, `HNew`, `HWNew`, `WNew`
      Increment successful realisations
    **end if**
  **end while**
  **Process Results:**
  Compute mean $S$, $I$, $R$, `HNew`, `HWNew`, `WNew`
  Compute confidence intervals for $I$, infections
**end for**
**Return:** `Results` with time-series and infection sources

**Table 1.** Parameters description, baseline values, and their ranges. These parameters are selected to be illustrative of a COVID-like infection; however, they are intended as exemplar parameterisation rather than reflecting a specific pathogen and population.

| Parameter | Description | Baseline Value | Low Value | High Value |
|---|---|---|---|---|
| $\mu$ | per contact infection probability per day | 0.18 | 0 | 1 |
| $\gamma$ | recovery probability per day | 0.16 | 0.1 | 1 |
| $n_h$ | household cluster size | 2 | 1 | 5 |
| $n_w$ | work cluster size | 5 | 2 | 20 |
| $x_h$ | household external connections | 2 | 0.5 | 5 |
| $x_w$ | work external connections | 2 | 0.5 | 5 |
| $\alpha$ | movement probability | 0.72 | 0 | 1 |
| $c_h$ | average household contacts during home time | 1 | 1 | 4 |
| $c_w$ | average contacts at work | 3 | 1 | 5 |
| $\kappa$ | coupling constant | 0.6031 | constant | constant |
| $\nu_\tau$ | steepness parameter (contact duration) | 1/60 | constant | constant |
| $\nu_d$ | steepness parameter (contact distance) | 1/10 | constant | constant |
| $\tau$ | contact duration required (minutes) | 15 | 1 | 30 |
| $d$ | threshold contact distance required (metres) | 2 | 0 | 3 |
| $T_h$ | average household contact duration (minutes) | 15 | 0 | 120 |
| $T_w$ | average contact duration at work (minutes) | 15 | 0 | 120 |
| $D_h$ | average household contact proximity (metres) | 2 | 0 | 5 |
| $D_w$ | average contact proximity at work (metres) | 2 | 0 | 5 |

##### *1.3.1 Linearization of the Force of Infection*

Following similar approach in [3], the linearised form of equations (11)-13 equations are:

$$P_h(t) \approx C_h \mu \frac{I(t)}{N} \left(1 - e^{-\kappa \frac{n_h x_h}{n_h + 1}}\right), \tag{18}$$

$$P_{hw}(t) \approx C_h \mu \frac{I(t)}{N} \left(1 - e^{-\kappa \frac{(1-\alpha) n_h x_h}{(1-\alpha) n_h + 1}}\right), \tag{19}$$

$$P_w(t) \approx C_w \mu \frac{I(t)}{N} \left(1 - e^{-\kappa \frac{\alpha n_w x_w}{\alpha n_w + 1}}\right). \tag{20}$$

Thus, the total force of infection, assuming that as $t \to 0$, then $1-P_h(t) \to 1$ as $P_h(0) \approx 0$ in equation (10) becomes:

$$\Delta(t) \approx \mu \frac{I(t)}{N}\left[C_h\left(1-e^{-\kappa\frac{n_h x_h}{n_h+1}}\right)+(1-\alpha)C_h\left(1-e^{-\kappa\frac{(1-\alpha)n_h x_h}{(1-\alpha)n_h+1}}\right)+\alpha C_w\left(1-e^{-\kappa\frac{\alpha n_w x_w}{\alpha n_w+1}}\right)\right]. \tag{21}$$

Define the effective transmission rate:

$$\beta_{\text{eff}} = \mu\left[C_h\left(1-e^{-\kappa\frac{n_h x_h}{n_h+1}}\right)+(1-\alpha)C_h\left(1-e^{-\kappa\frac{(1-\alpha)n_h x_h}{(1-\alpha)n_h+1}}\right)+\alpha C_w\left(1-e^{-\kappa\frac{\alpha n_w x_w}{\alpha n_w+1}}\right)\right], \tag{22}$$

so that:

$$\Delta(t) \approx \beta_{\text{eff}}\frac{I(t)}{N}. \tag{23}$$

#### 1.3.2 Transmission and Transition Matrices

The transmission term is:

$$\mathscr{F} = S(t)\Delta(t) \approx S(t)\beta_{\text{eff}}\frac{I(t)}{N}. \tag{24}$$

The transition term is:

$$\mathscr{T} = (1-\gamma)I(t). \tag{25}$$

The Jacobian matrices are:

$$F = \frac{\partial \mathscr{F}}{\partial I} = S(t)\frac{\beta_{\text{eff}}}{N}, \tag{26}$$

$$T = \frac{\partial \mathscr{T}}{\partial I} = 1-\gamma. \tag{27}$$

Since $0<\gamma<1$, $\rho(T) = |1-\gamma| < 1$. Then:

$$\mathbb{I}-T = \gamma, \quad [\mathbb{I}-T]^{-1} = \frac{1}{\gamma}. \tag{28}$$

The NGM is:

$$K = F[\mathbb{I}-T]^{-1} = \frac{S(t)\beta_{\text{eff}}}{\gamma N}. \tag{29}$$

The effective reproduction number is:

$$\mathscr{R}_t = \frac{S(t)\beta_{\text{eff}}}{\gamma N}. \tag{30}$$

Evaluate at disease-free equilibrium ($S(0) = N, I(0) = 0$), so the basic reproduction number is:

$$\mathscr{R}_0 = \frac{\beta_{\text{eff}}}{\gamma} = \frac{\mu}{\gamma}\left[C_h\left(1-e^{-\kappa\frac{n_h x_h}{n_h+1}}\right)+(1-\alpha)C_h\left(1-e^{-\kappa\frac{(1-\alpha)n_h x_h}{(1-\alpha)n_h+1}}\right)+\alpha C_w\left(1-e^{-\kappa\frac{\alpha n_w x_w}{\alpha n_w+1}}\right)\right]. \tag{31}$$

Substituting for $C_h$, and $C_w$:

$$C_h = \frac{4c_h}{\left(1+e^{\nu_\tau(\tau-T_h)}\right)\left(1+e^{-\nu_d(d-D_h)}\right)}, \tag{32}$$

$$C_w = \frac{4c_w}{\left(1+e^{\nu_\tau(\tau-T_w)}\right)\left(1+e^{-\nu_d(d-D_w)}\right)} \tag{33}$$

gives

$$\mathscr{R}_0 = \frac{4\mu}{\gamma}\Bigg[\frac{c_h\left[\left(1-e^{-\kappa\frac{n_hx_h}{n_h+1}}\right)+(1-\alpha)\left(1-e^{-\kappa\frac{(1-\alpha)n_hx_h}{(1-\alpha)n_h+1}}\right)\right]}{\left(1+e^{\nu_\tau(\tau-T_h)}\right)\left(1+e^{-\nu_d(d-D_h)}\right)} + \frac{\alpha c_w\left(1-e^{-\kappa\frac{\alpha n_wx_w}{\alpha n_w+1}}\right)}{\left(1+e^{\nu_\tau(\tau-T_w)}\right)\left(1+e^{-\nu_d(d-D_w)}\right)}\Bigg]. \tag{34}$$

#### 1.4 Sensitivity analysis of $\mathscr{R}_0$

The sensitivity analysis of $\mathscr{R}_0$ is presented in this section to deduce the impact of each parameter on $\mathscr{R}_0$. The local sensitivity analysis, which is a one-at-a-time (OAT) technique to find the influence of each parameter while keeping other parameters fixed [6], uses computation of partial derivatives of $\mathscr{R}_0$ with respect to each parameter $y_i$, ($\frac{\partial\mathscr{R}_0}{\partial y_i}$), to compute the sensitivity indices using the relationship

$$S_{y_i} = \frac{\partial\mathscr{R}_0}{\partial y_i}\frac{y_i}{\mathscr{R}_0}, \tag{35}$$

which is the normalised sensitivity index, which expresses the relative change in $\mathscr{R}_0$ due to a relative change in $y_i$.

#### 1.5 Partial Derivatives of $\mathscr{R}_0$

The partial derivatives of $\mathscr{R}_0$ (equation (34)) with respect to the parameters are as follows:

$$\frac{\partial \mathscr{R}_0}{\partial \mu} = \frac{4}{\gamma}\Bigg[\frac{c_h\left(1-e^{-\kappa\frac{n_h x_h}{n_h+1}}\right)}{\left(1+e^{\nu_\tau(\tau-T_h)}\right)\left(1+e^{-\nu_d(d-D_h)}\right)} + \frac{(1-\alpha)c_h\left(1-e^{-\kappa\frac{(1-\alpha)n_h x_h}{(1-\alpha)n_h+1}}\right)}{\left(1+e^{\nu_\tau(\tau-T_h)}\right)\left(1+e^{-\nu_d(d-D_h)}\right)} + \frac{\alpha c_w\left(1-e^{-\kappa\frac{\alpha n_w x_w}{\alpha n_w+1}}\right)}{\left(1+e^{\nu_\tau(\tau-T_w)}\right)\left(1+e^{-\nu_d(d-D_w)}\right)}\Bigg], \tag{36}$$

$$\frac{\partial \mathscr{R}_0}{\partial \gamma} = -\frac{4\mu}{\gamma^2}\Bigg[\frac{c_h\left(1-e^{-\kappa\frac{n_h x_h}{n_h+1}}\right)}{\left(1+e^{\nu_\tau(\tau-T_h)}\right)\left(1+e^{-\nu_d(d-D_h)}\right)} + \frac{(1-\alpha)c_h\left(1-e^{-\kappa\frac{(1-\alpha)n_h x_h}{(1-\alpha)n_h+1}}\right)}{\left(1+e^{\nu_\tau(\tau-T_h)}\right)\left(1+e^{-\nu_d(d-D_h)}\right)} + \frac{\alpha c_w\left(1-e^{-\kappa\frac{\alpha n_w x_w}{\alpha n_w+1}}\right)}{\left(1+e^{\nu_\tau(\tau-T_w)}\right)\left(1+e^{-\nu_d(d-D_w)}\right)}\Bigg], \tag{37}$$

$$\frac{\partial \mathscr{R}_0}{\partial c_h} = \frac{4\mu\left(\left(1-e^{-\kappa\frac{n_h x_h}{n_h+1}}\right)+(1-\alpha)\left(1-e^{-\kappa\frac{(1-\alpha)n_h x_h}{(1-\alpha)n_h+1}}\right)\right)}{\gamma\left(1+e^{\nu_\tau(\tau-T_h)}\right)\left(1+e^{-\nu_d(d-D_h)}\right)}. \tag{38}$$

$$\frac{\partial \mathscr{R}_0}{\partial c_w} = \frac{4\mu\alpha\left(1-e^{-\kappa\frac{\alpha n_w x_w}{\alpha n_w+1}}\right)}{\gamma\left(1+e^{\nu_\tau(\tau-T_w)}\right)\left(1+e^{-\nu_d(d-D_w)}\right)}, \tag{39}$$

$$\frac{\partial \mathscr{R}_0}{\partial \alpha} = \frac{4\mu}{\gamma}\Bigg\{\frac{c_h\left[-\left(1-e^{-\kappa\frac{(1-\alpha)n_h x_h}{(1-\alpha)n_h+1}}\right)-(1-\alpha)\kappa\frac{n_h x_h e^{-\kappa\frac{(1-\alpha)n_h x_h}{(1-\alpha)n_h+1}}}{((1-\alpha)n_h+1)^2}\right]}{\left(1+e^{\nu_\tau(\tau-T_h)}\right)\left(1+e^{-\nu_d(d-D_h)}\right)} \tag{40}$$

$$+ \frac{c_w}{\left(1+e^{\nu_\tau(\tau-T_w)}\right)\left(1+e^{-\nu_d(d-D_w)}\right)}\left[\left(1-e^{-\kappa\frac{\alpha n_w x_w}{\alpha n_w+1}}\right)+\alpha\kappa\frac{n_w x_w e^{-\kappa\frac{\alpha n_w x_w}{\alpha n_w+1}}}{(\alpha n_w+1)^2}\right]\Bigg\}, \tag{41}$$

$$\frac{\partial \mathscr{R}_0}{\partial n_h} = \frac{4\mu c_h \kappa x_h}{\gamma\left(1+e^{\nu_\tau(\tau-T_h)}\right)\left(1+e^{-\nu_d(d-D_h)}\right)}\left[\frac{e^{-\kappa\frac{n_h x_h}{n_h+1}}}{(n_h+1)^2}+\frac{(1-\alpha)^2 e^{-\kappa\frac{(1-\alpha)n_h x_h}{(1-\alpha)n_h+1}}}{((1-\alpha)n_h+1)^2}\right], \tag{42}$$

$$\frac{\partial \mathscr{R}_0}{\partial n_w} = \frac{4\mu\alpha^2 c_w \kappa x_w e^{-\kappa\frac{\alpha n_w x_w}{\alpha n_w+1}}}{\gamma(\alpha n_w+1)^2\left(1+e^{\nu_\tau(\tau-T_w)}\right)\left(1+e^{-\nu_d(d-D_w)}\right)}, \tag{43}$$

$$\frac{\partial \mathscr{R}_0}{\partial x_h} = \frac{4\mu c_h \kappa}{\gamma}\left[\frac{n_h e^{-\kappa \frac{n_h x_h}{n_h+1}}}{(n_h+1)\left(1+e^{\nu_\tau(\tau-T_h)}\right)\left(1+e^{-\nu_d(d-D_h)}\right)} + \frac{(1-\alpha)^2 n_h e^{-\kappa \frac{(1-\alpha)n_h x_h}{(1-\alpha)n_h+1}}}{((1-\alpha)n_h+1)\left(1+e^{\nu_\tau(\tau-T_h)}\right)\left(1+e^{-\nu_d(d-D_h)}\right)}\right]. \tag{44}$$

$$\frac{\partial \mathscr{R}_0}{\partial x_w} = \frac{4\mu\alpha^2 c_w \kappa n_w e^{-\kappa \frac{\alpha n_w x_w}{\alpha n_w+1}}}{\gamma(\alpha n_w+1)\left(1+e^{\nu_\tau(\tau-T_w)}\right)\left(1+e^{-\nu_d(d-D_w)}\right)}, \tag{45}$$

$$\frac{\partial \mathscr{R}_0}{\partial \tau} = -\frac{4\mu\nu_\tau}{\gamma}\left[\frac{c_h\left(1-e^{-\kappa\frac{n_h x_h}{n_h+1}} + (1-\alpha)\left(1-e^{-\kappa\frac{(1-\alpha)n_h x_h}{(1-\alpha)n_h+1}}\right)\right)e^{\nu_\tau(\tau-T_h)}}{\left(1+e^{\nu_\tau(\tau-T_h)}\right)^2\left(1+e^{-\nu_d(d-D_h)}\right)} + \frac{\alpha c_w\left(1-e^{-\kappa\frac{\alpha n_w x_w}{\alpha n_w+1}}\right)e^{\nu_\tau(\tau-T_w)}}{\left(1+e^{\nu_\tau(\tau-T_w)}\right)^2\left(1+e^{-\nu_d(d-D_w)}\right)}\right], \tag{46}$$

$$\frac{\partial \mathscr{R}_0}{\partial d} = \frac{4\mu\nu_d}{\gamma}\left[\frac{c_h\left(1-e^{-\kappa\frac{n_h x_h}{n_h+1}} + (1-\alpha)\left(1-e^{-\kappa\frac{(1-\alpha)n_h x_h}{(1-\alpha)n_h+1}}\right)\right)e^{-\nu_d(d-D_h)}}{\left(1+e^{\nu_\tau(\tau-T_h)}\right)\left(1+e^{-\nu_d(d-D_h)}\right)^2} + \frac{\alpha c_w\left(1-e^{-\kappa\frac{\alpha n_w x_w}{\alpha n_w+1}}\right)e^{-\nu_d(d-D_w)}}{\left(1+e^{\nu_\tau(\tau-T_w)}\right)\left(1+e^{-\nu_d(d-D_w)}\right)^2}\right], \tag{47}$$

$$\frac{\partial \mathscr{R}_0}{\partial T_h} = \frac{4\mu\nu_\tau}{\gamma}\left[\frac{c_h\left(1-e^{-\kappa\frac{n_h x_h}{n_h+1}} + (1-\alpha)\left(1-e^{-\kappa\frac{(1-\alpha)n_h x_h}{(1-\alpha)n_h+1}}\right)\right)e^{\nu_\tau(\tau-T_h)}}{\left(1+e^{\nu_\tau(\tau-T_h)}\right)^2\left(1+e^{-\nu_d(d-D_h)}\right)}\right], \tag{48}$$

$$\frac{\partial \mathscr{R}_0}{\partial T_w} = \frac{4\mu\nu_\tau\alpha c_w\left(1-e^{-\kappa\frac{\alpha n_w x_w}{\alpha n_w+1}}\right)e^{\nu_\tau(\tau-T_w)}}{\gamma\left(1+e^{\nu_\tau(\tau-T_w)}\right)^2\left(1+e^{-\nu_d(d-D_w)}\right)}, \tag{49}$$

$$\frac{\partial \mathscr{R}_0}{\partial D_h} = -\frac{4\mu\nu_d}{\gamma}\left[\frac{c_h\left(1-e^{-\kappa\frac{n_h x_h}{n_h+1}} + (1-\alpha)\left(1-e^{-\kappa\frac{(1-\alpha)n_h x_h}{(1-\alpha)n_h+1}}\right)\right)e^{-\nu_d(d-D_h)}}{\left(1+e^{\nu_\tau(\tau-T_h)}\right)\left(1+e^{-\nu_d(d-D_h)}\right)^2}\right], \tag{50}$$

$$\frac{\partial \mathscr{R}_0}{\partial D_w} = -\frac{4\mu\nu_d}{\gamma}\left[\frac{\alpha c_w\left(1-e^{-\kappa\frac{\alpha n_w x_w}{\alpha n_w+1}}\right)e^{-\nu_d(d-D_w)}}{\left(1+e^{\nu_\tau(\tau-T_w)}\right)\left(1+e^{-\nu_d(d-D_w)}\right)^2}\right]. \tag{51}$$

From the sensitivity indices, a positive $S_{y_i}$ signifies that the function value (in this model, $\mathscr{R}_0$) will increase as the parameter value $y_i$ increases, while a negative $S_{y_i}$ means that the function value will decrease as the parameter $y_i$ increases [7]. The analytical expressions of the partial derivatives computed above do not communicate to us the order of importance in the sensitivity indices of the parameters given their default values and ranges; as such, numerical results are presented in the next section.

### 1.6 Numerical simulation

The Sobol Global Sensitivity Analysis method [8, 9] was used. In this approach, the steps in **Algorithm Box 7** were followed to implement the sensitivity analysis in MATLAB:

**Algorithm 7** Sobol Sensitivity Analysis Algorithm

**Input:** Parameter bounds (Table 1), number of samples $N$, function evaluating $\mathscr{R}_0$ (basic reproduction number), parameters $X_i$.
**Output:** First-order Sobol indices $S_i$, total-order Sobol indices $S_{T_i}$ for each parameter $i$.
**Initialize:**
Specify lower and upper bounds for each parameter $X_i$.
Generate two independent Latin Hypercube Sampling (LHS) sets $A$ and $B$, each an $N \times k$ matrix where each row is a sample of all $k$ parameters $X_i$ and each column contains $N$ samples for a single parameter, drawn within its bounds.
**Model Evaluation:**
**for** each sample $a \in A$ and $b \in B$ **do**
    Compute ${\mathscr{R}_0}^{(a)}$ and ${\mathscr{R}_0}^{(b)}$ using the function that evaluates $\mathscr{R}_0$.
**end for**
**Mixed Sample Evaluation:**
**for** each parameter $i$ **do**
    Construct mixed sample set $C$ by replacing $i$-th column of $A$ with $i$-th column of $B$.
    Compute ${\mathscr{R}_0}^{(c)}$ for each sample $c \in C$ using the function that evaluates $\mathscr{R}_0$.
**end for**
**Variance Calculations:**
Compute total variance across set A: $V \leftarrow \mathrm{Var}({\mathscr{R}_0}^{(A)})$.
**for** each parameter $i$ **do**
    Compute first-order contribution: $V_i \leftarrow \mathrm{mean}({\mathscr{R}_0}^{(A)}{\mathscr{R}_0}^{(C)}) - \mathrm{mean}({\mathscr{R}_0}^{(A)})^2$.
    Compute total-order contribution: $V_{T_i} \leftarrow \frac{\mathrm{mean}(({\mathscr{R}_0}^{(A)} - {\mathscr{R}_0}^{(C)})^2)}{2}$.
**end for**
**Sobol Indices:**
**for** each parameter $i$ **do**
    Compute first-order Sobol index: $S_i \leftarrow \frac{V_i}{V}$.
    Compute total-order Sobol index: $S_{T_i} \leftarrow \frac{V_{T_i}}{V}$.
**end for**
**Return:** $S_i$, $S_{T_i}$ for each parameter $i$.

The total variance $V$ is calculated as:

$$V = \text{Var}(\mathscr{R}_0{}^{(A)}),$$

where $\mathscr{R}_0{}^{(A)}$ is the model output evaluated using the parameter samples for each $c \in C$. This is a measure of how "spread out" the model outputs are, which quantifies how much uncertainty exists in the model output due to the variation in all input parameters. The first-order variance contribution $V_i$ captures the variance in the output that is solely due to parameter $i$, without considering interactions with other parameters. It is calculated as:

$$V_i = \text{mean}(\mathscr{R}_0{}^{(A)}\mathscr{R}_0{}^{(C)}) - \text{mean}(\mathscr{R}_0{}^{(A)})^2.$$

Where $\mathscr{R}_0{}^{(A)}$ represents the model output when a set of parameters $A$ is used. $\mathscr{R}_0{}^{(C)}$ represents the model output when only parameter $i$ is replaced by its counterpart in set $B$, while all other parameters remain the same. The term $\text{mean}(\mathscr{R}_0{}^{(A)}\mathscr{R}_0{}^{(C)})$ captures how much the outputs of $A$ and $C$ are correlated due to parameter $i$. Subtracting $\text{mean}(\mathscr{R}_0{}^{(A)})^2$ removes the baseline variance that arises purely from the mean output value, leaving only the variance contribution from parameter $i$. The total-order variance contribution $V_{T_i}$ captures all the variance in the output that involves parameter $i$, including its interactions with other parameters. It is calculated as:

$$V_{T_i} = \frac{\text{mean}\left((\mathscr{R}_0{}^{(A)} - \mathscr{R}_0{}^{(C)})^2\right)}{2}.$$

Where $(\mathscr{R}_0{}^{(A)} - \mathscr{R}_0{}^{(C)})^2$ measures the squared difference in output due to changing parameter $i$. The mean of these squared differences represents how much variance is introduced by $i$, including any interactions $i$ has with other parameters. Dividing by 2 adjusts for the fact that the squared difference includes contributions in both directions (from $A$ to $C$ and vice versa).

### 2 Results

The analytical expression of the basic reproduction number derived using this framework was given in the methods section (equation (34)).

Figures 4 and **??** show that the EBM dynamics are well within a 95% prediction interval (PI) of IBM, and follow similar behaviour for increasing $x_h$ and $x_w$, although the peak infections for the EBM simulation are higher than the mean of the IBM simulation. These results are aimed at verifying whether the EBM developed here is a good approximation of the IBM.

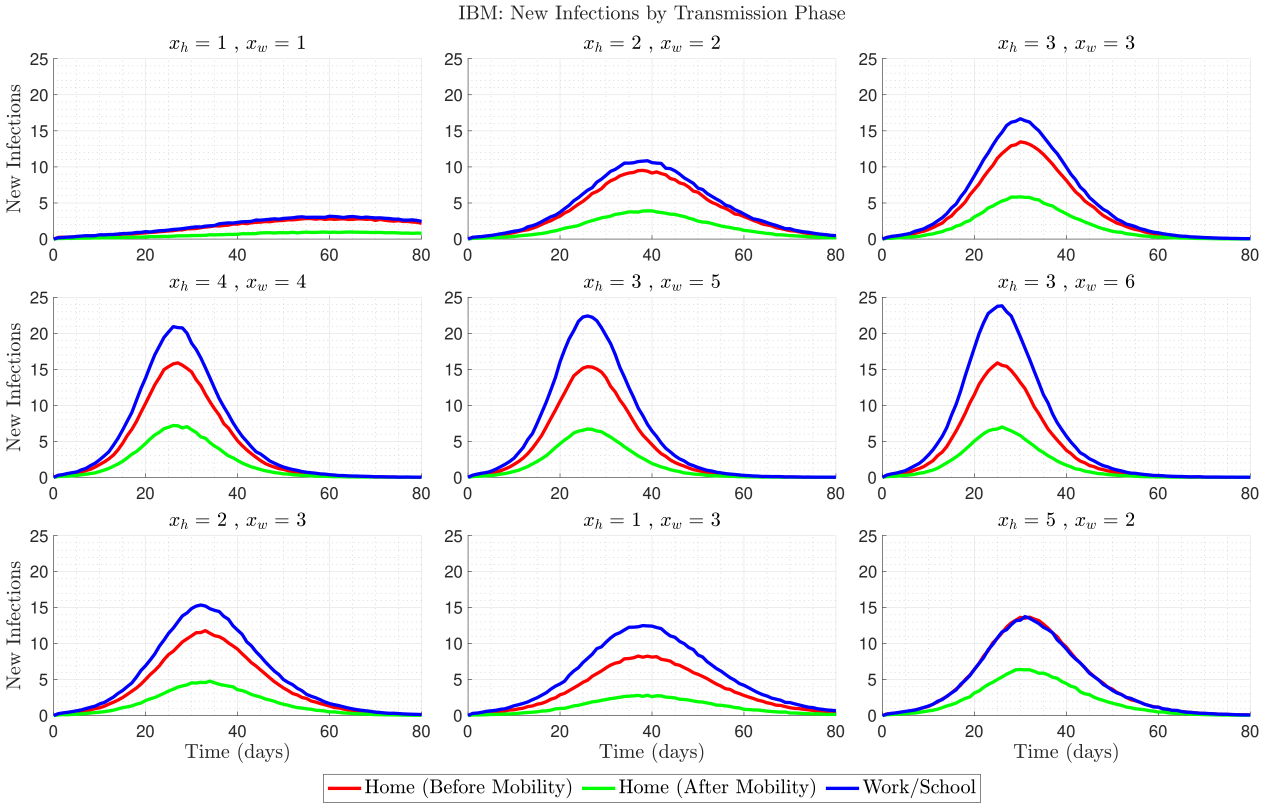


**Figure 3.** IBM simulation results showing daily new infections (average of 1,000 realisations) stratified by transmission phase: home before mobility (red), home after mobility (green), and workplace (blue).


### Acknowledgements

This research is fully funded by the Institute for Global Pandemic Planning, Warwick Medical School, University of Warwick, UK.


### Author contributions statement

Conceptualisation: M. L. S.
Methodology: M. L. S.
Supervision: A.S. & K. S. R.
Software: K. S. R. & M. L. S.
Visualisation: K S. R & M. L. S.
Writing – original draft: M. L. S.
Writing – review & editing: A. S., K. S. R. & M. L. S.

### Data availability

MATLAB code will be accessible via GitHub upon publication of the manuscript.

### Ethics declarations

This study focuses on the theoretical development of a mathematical model for the spread of a hypothetical infectious disease. No data was collected and analysed.

### Additional information

**Competing interests**. The authors declare no competing interests. For open access, the authors have applied a Creative Commons Attribution (CC-BY) licence to any Author Accepted Manuscript version arising from this submission.

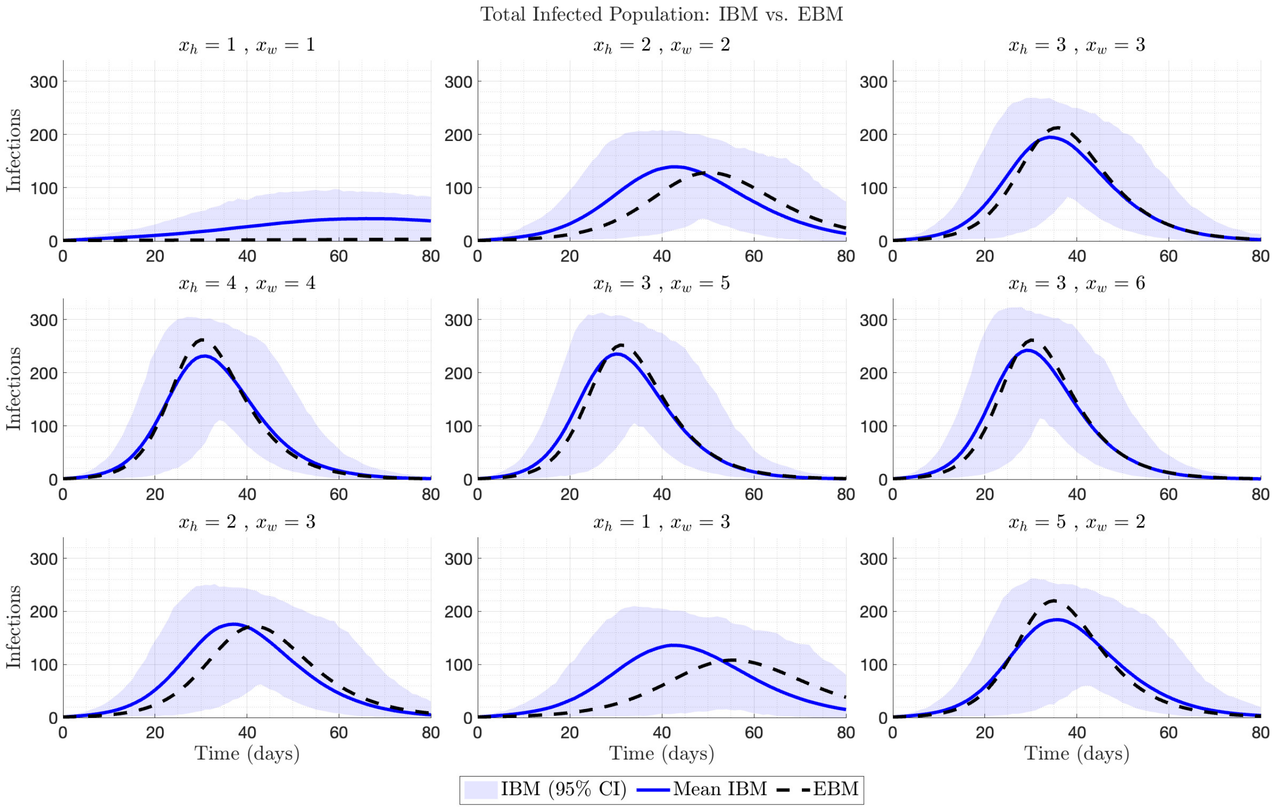


**Figure 4.** Comparison of total infected population dynamics between the individual-based model (IBM, shaded region showing 95% prediction interval-average of 1,000 realisations) and equation-based model (EBM, solid line) across different external connection configurations. Each subplot represents a specific number of external household ($x_h$) and workplace ($x_w$) connections, demonstrating how increased connectivity affects epidemic progression in both modelling approaches.
